# Modelling airborne transmission of SARS-CoV-2 using CARA: Risk assessment for enclosed spaces

**DOI:** 10.1101/2021.10.14.21264988

**Authors:** Andre Henriques, Nicolas Mounet, Luis Aleixo, Philip Elson, James Devine, Gabriella Azzopardi, Marco Andreini, Markus Rognlien, Nicola Tarocco, Julian Tang

## Abstract

The global crisis triggered by the COVID-19 pandemic has highlighted the need for a proper risk assessment of respiratory pathogens in indoor settings. This paper documents the COVID Airborne Risk Assessment (CARA) methodology, to assess the potential exposure of airborne SARS-CoV-2 viruses, with an emphasis on the effect of certain virological and immunological factors in the quantification of the risk. The proposed model is the result of a multidisciplinary approach linking physical, mechanical and biological domains, benchmarked with clinical and experimental data, enabling decision makers or facility managers to perform risk assessments against airborne transmission. The model was tested against two benchmark outbreaks, showing good agreement. The tool was also applied to several everyday-life settings, in particular for the cases of a shared office, classroom and ski cabin. We found that 20% of infected hosts can emit approximately 2 orders of magnitude more viral-containing particles, suggesting the importance of *super-emitters* in airborne transmission. The use of surgical-type masks provides a 5-fold reduction in viral emissions. Natural ventilation through the opening of windows at all times are effective strategies to decrease the concentration of virions and slightly opening a window in the winter has approximately the same effect as a full window opening during the summer. Although vaccination is an effective protection measure, non-pharmaceutical interventions, which significantly reduce the viral density in the air (ventilation, masks), should be actively supported and included early in the risk assessment process. We propose a critical threshold value approach which could be used to define an acceptable risk level in a given indoor setting.

## 1 Introduction

Currently, the existing public health measures point to the importance of proper building and environmental engineering control measures, such as proper Indoor Air Quality (IAQ). The COVID-19 pandemic has raised increased awareness on airborne transmission of respiratory viruses in indoor settings. Of the main modes of viral transmission, the airborne route of SARS-CoV-2 seems to have a significant importance to the spread of COVID-19 infections world-wide [1]. Furthermore, infection through aerosol inhalation could lead to more severe disease than infection from fomites [2]. The potential for presymptomatic and asymptomatic transmission is also reported, with evidence suggesting that 30-70% of transmission happens before symptom onset [3], with viral loads peaking at around the time of symptom onset. This contrasts with other coronaviruses which peak at around 7-14 days after symptom onset [4]. The high viral loads around symptom onset suggest that SARS-CoV-2 could be easily transmissible at an early stage of infection.

Facility managers are facing a new paradigm where the need for a concrete, quick and simplified tool to prevent airborne transmission in buildings and other enclosed spaces is becoming an essential part of any occupational health and safety risk assessment.

In occupational health and safety, the best way to ensure proper protection is to fully understand i) what are the causes of a given hazardous event and how to prevent it; ii) what are consequences arising from the hazardous event and how to protect from it. For any given risk, in order to consider the appropriate mitigation and risk control measures for indoor spaces (e.g. workplaces, household, public spaces), a multidisciplinary risk-based approach is essential.

In this paper, a physical model is proposed, adapted from previous implementations of other infectious models such as the Wells-Riley approach [5, 6], to simulate the concentration of infectious viruses in an enclosed indoor volume, wherein infectious occupants with COVID-19 are shedding SARS-CoV-2 viruses. The present study focuses on the so-called ‘long-range’ airborne transmission route, assuming a well-mixed box model with a homogeneous viral concentration in the entire volume and that occupants in the room are physically distant from each other. The model follows a probabilistic approach to deal with the uncertainties tied to the concerned variables, such as the characteristics of this novel virus, including the properties of the emerging Variants of Concern (VOC). Various aspects of the medical, biological, mechanical and physical characteristics of the respiratory airborne pathogens are taken into account, in particular the mechanistic process of respiratory droplet nuclei emission, the effectiveness of face covering, the dilution with outdoor air, the impact of particulate filtration, the inactivation of infectious viruses, host immunity and dispersion models in indoor environments.

Human presence is the generation source of expiratory droplets and droplet nuclei potentially containing virions, when performing vocal or pure respiratory activities [7, 8]. The droplets and droplet nuclei which are sufficiently small to be aerodynamically suspended in the air may be inhaled by the exposed occupants in the same indoor setting. Particular attention is given to the winter period as a result of increased indoor gatherings with limited air renewal, resulting in increased exposure duration [9], and the low relative humidity of the indoor ambient air, mainly due to the effect of central heating (e.g. via superheated water radiators). Low humidity air increases the evaporation of droplets and consequently the number of airborne droplet nuclei [10]. This increases the virus survival in air and reduces immune defenses of the exposed hosts [11].

The methodology presented in this paper exhibits five main aspects: 1) the generation (or emission) rate of viruses coming from the infected host(s), which is a result of the respiratory activity combined with the possible use of face covering (source control); 2) the indoor viral removal rate resulting from ventilation or air filtration, viability decay and gravitational settlement; 3) the indoor viral concentration profile over time resulting from the balance of the two previous quantities; 4) the accumulated viral dose absorbed by an exposed host and deposited in the respiratory tract; and 5) the probability of infection (or transmission) resulting from such a dose.

To estimate the generation rate, a probabilistic approach is considered by parameterising the viral load distributions in the respiratory tract and the volumetric concentration of droplets emitted, measured for different activities [12]. By convention, droplets and droplet nuclei will be discussed in this paper as ‘airborne particles’. Once an infected occupant is emitting viruses into the volume, the physical and mechanical behavior of the floating virus-containing particles is simulated, based on the assumption of a homogeneous mixture in a finite volume. With a realistic set of inputs, including a complex ventilation algorithm with openable windows and a flexible occupancy profile, the model allows for a combination and comparison of various mitigation measures, aimed at properly assessing the situation tailored to common practices in indoor settings. The main goal of this paper is to improve the common understanding in the modelling of airborne transmission and identify the pivotal parameters, in order to develop a quantitative action plan to help building engineers, facility managers and household individuals, in identifying which measures or combination of measures are most suitable, allowing for a tailored risk assessment and targeted investment. The results are compared with similar studies from literature.

The model, its assumptions and data used will be presented in Section 2. The set of possible occupation and activity profiles taken into account in the model are listed in Section 3, and simulation results in various situations follow in Section 4. These are then discussed in Section 5, and our conclusions are finally presented in Section 6.

## 2 Methods

The infection model presented in this paper, is computed using an Open Source software called the COVID Airborne Risk Assessment (CARA). The availability statement of the software and associated data is mentioned in Section 6. The methodology behind the infection model is split into five modules: Source (emission), Dispersion (removal rate), Exposure (concentration), Dose (inhalation) and Risk (infection).

Several risk assessment tools have opted to adapt the notion of ‘quantum of infection’, introduced in the 1950’s by W. F. Wells, suggesting a hypothetical infectious dose unit for a certain pathogen [13]. The *quanta* is generally estimated from epidemiological data following an outbreak investigation, using a reverse engineering approach predicting the environmental conditions at the time of the outbreak. In the framework of a risk management and risk prevention approach, we opted to study the projection of possible new infections before the potential outbreak takes place. Thus, we decided to adopt an approach by relating the physiological mechanism of respiratory particle emissions, introduced by Nicas et al. [5] for Tuberculosis and adapted by Buonanno et al. [12] for COVID-19, while integrating the virological and pathological characteristics of such respiratory pathogens. The model assumes a homogeneous viral distribution in the respiratory tract [14] and that the number of virions in a given particle is proportional to its size, as well as a homogeneous mixture of airborne particles in a given room volume (i.e. well-mixed box model). Several authors have characterized the amount of respiratory particles and their size distribution, during different vocal activities [7, 15–17]. The underlying question relates to the relationship between a mechanistic approach and the virological aspects, which we propose to address in this paper. Since typical coronaviruses seem to require more than one pathogen to initiate infection [18, 19], this study includes the concept of an equivalent infectious dose ID. According to Sze et al. [20], the dose-response models for such risk assessment are more precise, as many influencing factors can be determined explicitly, allowing for fewer implicit errors in general. On the other hand, such models are not yet available for SARS-CoV-2, hence a solution is described in this paper, taking into account the effect of host immunity.

Many of the model variables (such as emission rate, removal rate, and concentration) are considered for a given aerosol diameter *D*, as the dynamics in the room and the deposition efficiency in the respiratory tract, depends on the particle size. The resulting dose is then computed over a distribution of particle diameters, which is then followed by a Monte Carlo integration. Furthermore, some other variables (such as viral load, infectious dose, mask inward efficiency, breathing rate) are treated as random to account for their aleatory uncertainties; the related distribution descriptors are defined by data available in the literature. Finally, the probability of infection is estimated by using plain Monte Carlo sampling algorithms.

### 2.1 Emission rate (vR)

The emission rate of virus per unit diameter, vR (in virion h^-1^μm^-1^), is estimated by considering the volumetric emissions of respiratory particles of a given diameter *D* by the infected host(s), and the virological characteristics*. The former properties are evaluated for three different expiratory activities: i) Breathing (*index ‘*b*’*); ii) Speaking (*index ‘*sp*’*), iii) Shouting/Singing (*index ‘*sh*’*). While performing the expiratory activities, the particle emissions are known to be affected by the physical activity and loudness of the infected host’s vocalisation, since the faster one breathes or the louder one speaks, the more particles per unit volume are being emitted [7, 16]. Subsequently the emission rate includes the contribution of tidal volumes and amplitude of the voice. The virological characteristics include the density of viral copies from nasopharyngeal (NP) swabs. The viral emission rate can be calculated using the following formulation:

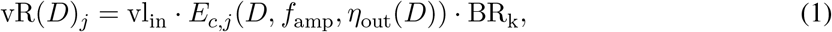

where vl_in_ is the viral load inside the infected host’s respiratory tract (in RNA copies per mL); *E*_*c,j*_ represents the volumetric particle emission concentration per unit diameter (in mL m^-3^μm^-1^), for a given expiratory activity *j* and as a function of the vocalisation amplification factor *f*_amp_ and the outward mask efficiency *η*_out_(*D*) (which also depends on the particle size); BR_k_ (in m^3^ h^-1^) is the breathing flow rate for a given physical activity *k*. For the purposes of this model, we will assume a 1:1 ratio between RNA copies and virions when interpreting PCR viral load results, though for PCR assays targeting the N (nucleocapsid) gene, this ratio may far exceed 1. The volume of the respiratory particles are calculated assuming each is a perfect sphere. In the present model, vR is assumed to be piecewise constant over time, for each type of vocalisation/respiratory activity, and is considered constant when the infected host is present (vR = 0 when the infected host is absent).

Due to the large variability of the different variables discussed in this paper, a probabilistic approach is used to determine vR. The methods and variables are described in Sections 2.1.1 -2.1.4 and summarized in Section 2.5.2.

Note that in the following subsections, for the purpose of simplification, we will omit the index *j* for vR and the quantities that depend on it, as each type of expiratory activity will be computed separately.

#### 2.1.1 *Viral Load (*vl)

In this paper, we describe the viral load as two separate parameters: the viral load inside the infected host, vl_in_, determined by RT-PCR assays from nasopharyngeal (NP) swabs, and the viral load outside the infected host, vl_out_, defined as the number of respiratory particles emitted from the mouth or nose during a given time *t* (in hours):

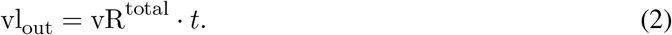

For vl_in_, data shows a large variability, ranging from 10^2^–10^11^ RNA copies per mL, covering symptomatic, presymptomatic and asymptomatic persons [21–23]. The large variability in these viral load values are related to the high dynamic range over the course of the infection and would largely impact vR. This aspect is particularly relevant when the uncertainty lies on the virological conditions of the infected host during transmission. Hence, here we considered statistical distributions where the baseline descriptors correspond respectively to a mean (SD) of 6.6 (1.7) log_10_ RNA copies per mL. These values were determined from the available dataset of approximately 20000 RT-PCR assays, sampled from February to April 2020 [23]. This parameter is a variable in our model, which could be adjusted to take into account new datasets.

In December 2020, an increasing portion of cases caused by a more transmissible new variant (Alpha) was observed in the United Kingdom. This has since been superseded by several other ‘Variants Of Concern’ (VOC) reported to have a significant effect on the risk of transmitting SARS-CoV-2. The precise mechanism(s) for increased transmissibility are not yet definitively understood at the time of writing. While preliminary laboratory results indicated evidence for higher viral load in infected individuals [24], it now appears that these elevated values were not substantiated by more recent studies [25]. This observation is consistent with the dynamics of cycle threshold (Ct) scores seen during rapid growth phases of the epidemic, as reported in Ref. [26]. There is now emerging evidence to indicate that the Delta VOC may combine both increased infectiousness and observably higher average viral loads compared to the wild type strains or the Alpha VOC, although the statistical significance of increases in viral load remains to be conclusively demonstrated.

#### 2.1.2 *Expiratory particle emissions (*E_c,j_)

During different vocalisation activities, or by simply breathing, a large amount of particles are emitted from the mouth and/or nose, originating from the respiratory tract [27, 28]. Particles of diameter smaller than 100 μm are likely to become airborne and can remain suspended in the air from seconds to hours, because of their reduced size and settling velocity compared to larger droplets [29]. Data on experimental studies measured the aerosolized particle concentration and size distribution [7, 30], although the aerosol sampling mechanisms employed, e.g. aerodynamic particle sizer (APS) or optical particle counter (OPC), are generally not capable of measuring the diameter of the respiratory droplets prior to evaporation [15], which occurs quasi-instantaneously after leaving the mouth or nose [31]. Understanding the initial diameter of the particle, prior to evaporation, is crucial for the quantification of the volumetric emissions [12] and consequently the emission rate (vR).

The particle emissions and their size distribution vary depending on the vocalisation activity. Johnson et al. [15] studied the size distributions of particle emissions for different expiratory activities and found three distinct modes associated with different anatomical processes in the respiratory tract: one originating from the bronchial region while breathing, another near the larynx (housing the vocal cords) which is highly active while speaking and singing; and one from the oral cavity (i.e. mouth) which is active during any vocalisation. The volumetric particle emission concentration (*E*_*c,j*_) is, therefore, modeled according to the aforementioned paper using a tri-modal log-normal distribution model (BLO model) [15], weighted by the particle size distribution multiplied by the volume at a given diameter (assuming each particle is a perfect sphere). In the same reference, the author included an evaporation factor (*f*_evap_) of 0.5 to take into account the ratio between desiccated and saturated particles. Here, we propose to use an evaporation factor of 0.3, based on more recent studies [32], considering an average protein content between 3 and 76 mg per mL of nasal fluid.

The particle emissions of the larynx (L) and oral (O) modes are equally affected by the amplitude of the vocalisation (i.e. loudness) and this relationship is found to scale linearly, while maintaining a constant size distribution [16]. An amplification factor (*f*_amp_) is used to scale the emission concentration relative to ‘Speaking’, which is used during vocalisation. The bronchial (B) mode, on the other hand, is not affected by the amplitude of vocalisation, hence the emission concentration during ‘Breathing’ is not impacted by this effect since only the B mode is active.

To make use of this data for the purpose of our study, we need to compute the volume of respiratory fluid emitted by the host, per volume of exhaled air, per unit diameter, *E*_*c,j*_ (*D*), in mL m^-3^μm^-1^, and for a given diameter *D*. This is given by

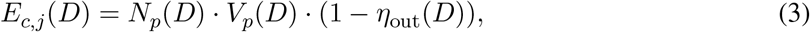

where *N*_*p*_(*D*) is the number of particles of this size (depending on the expiratory activity through *f*_amp_, see below), *V*_*p*_(*D*) is their individual volume and *η*_out_(*D*) is the outward mask efficiency (cf. Section 2.1.3) for this diameter. Based on the BLO model, the number of emitted particles of a given size can be obtained from

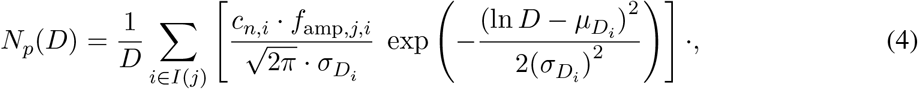

where *I*(*j*) is a subset of {B, L, O} determined by the expiratory activity *j*: for breathing *I*(b) = {B}, for speaking or shouting *I*(sp) = *I*(sh) = {B, L, O}; 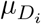 and 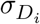 are the mean and standard deviation of the natural logarithm of the diameter for each mode (in ln μm); *c*_*n,i*_ is the total particle emission concentration for each mode. The amplification factor *f*_amp,*j*_ (*i*) follows [16]:

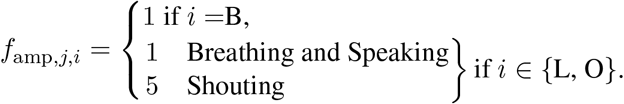

Table 1 provides the list of variables and the related distribution descriptors adopted to compute *E*_*c,j*_ from Eqs. (3) and (4) (the size distribution particle emission concentrations can be found in Supplementary Fig S.1). Particle emission concentration have been extensively reported by Bourouiba [33] with values ranging from approximately 0.01 to 4 particles cm^-3^ for breathing and 0.01 to 1 particles cm^-3^ for speaking. A parametric study was conducted to produce a fit with experimental data for viral emission rates (cf. Section 4).

**Table 1:** Parameters of the BLO model used in the volumetric particle emission concentration in Eq. (3). The geometric mean (GM) and geometric standard deviation (GSD) of the log-normal distributions for the particle diameters are also shown. Values for *c*_*n,i*_ are taken from Ref. [7] and particle diameter distribution parameters are extracted from Ref. [15], applying an evaporation correction factor *f*_evap_ = 0.3.

| Tri-modal parameters for $E_{c,j}$<sup>a</sup> | | | | | |
| --- | --- | --- | --- | --- | --- |
| Mode (*i*) | $c_{n,i} [\text{cm}^{-3}]$ | $\mu_{D_i} [\ln \mu\text{m}]$ | GM [$\mu\text{m}$] | $\sigma_{D_i} [\ln \mu\text{m}]$ | GSD [$\mu\text{m}$] |
| B | 0.06<sup>b</sup> | 0.99 | 2.69 | 0.26 | 1.30 |
| L | 0.2<sup>b</sup> | 1.39 | 4.01 | 0.51 | 1.67 |
| O | 0.001 | 4.98 | 145.5 | 0.59 | 1.80 |

<sup>a</sup> Unit conversion will be necessary to compute $E_{c,j}$ .
<sup>b</sup> Obtained following a parametric study (see Section 4).

To obtain the total expiratory emission in mL m^-3^, one has to integrate Eq. (3) over all particle diameters:

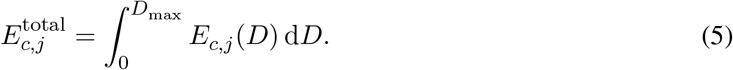

Since this paper is focused on airborne transmission, the limits of integration are set from 0 to *D*_max_ = 30 μm, which correspond to a desiccated particle diameter up to roughly 10 μm. This assumption is reasoned knowing that respiratory aerosols up to 10 μm (i.e. PM10) represent 99% of the total emission concentration while speaking [15].

Note that the integral in Eq. (5), used to compute the total emission, is performed using a Monte Carlo sampling of the particle diameters which follow the distribution given by *N*_*p*_(*D*) in Eq. (4). In the model, the integral is actually not performed at this stage but later when computing the dose, since other parameters also depends on *D* (see Section 2.4).

#### 2.1.3 Outward effect of face covering (η_out_)

Face coverings are reported to be a very efficient source control measure against infection prevention and disease control [34–39]. According to the basic prevention principle of risk assessments, reducing the hazard at the source is at the top of the priority list in terms of mitigation measures. The so-called surgical masks are widely used and recognised as appropriate face covering devices for source control. These masks are manufactured following strict performance and quality requirements and are certified by the applicable national authorities. The minimum material filtration efficiency accepted in, e.g., the USA and European Union is 95 %, using the test standards ASTM F2101 and EN 14683 [40], respectively. It is important to note that these results are the filtration efficiencies of the material and do not consider the losses due to the actual positioning on the wearer’s face, namely leakage. Since surgical masks are not meant to act as personal protective equipment (PPE) such as N95 or FFP2 masks, there are no requirements for leak-tightness in the test standards mentioned above. Both standards use a mean particle size of 3.0 *±* 0.3 μm for the measurements, although, when breathing, the majority of the emitted particles are smaller than 3 μm [7], even when considering a saturated particle size with a geometrical mean of 2.7 μm (cf. Table 1). Reducing the particles size will have an effect on the filtration efficiency. Recent studies measured the outward filtration efficiency for surgical masks of 80% while breathing [38] and 60−75% in the 0.7−2 μm size range [41]. The result of these measurements include the effect of leaks, without performing any fittest or fit-check procedure. The certification requirement of 95% efficiency may be used for particle sizes ≥ 3 μm, corrected to take into account the leakages. We assume a total leakage of 15% through the sides, nose and chin [42], which would yield an equivalent outward efficiency of 82% at sizes larger than 3 *≥* μm, comparable to the same measurements [38, 41]. In this paper, we use the values for outward efficiency (*η*_out_) of surgical masks, as a function of the particle diameter (Supplementary Fig. S.2).

The use of Personnel Protective Equipment - PPE (e.g. respirators), such as N95 and FFP2, are found to have a similar effect in terms of source control [38] and thus are assumed here to have an equivalent outward efficiency. *η*_out_ is equal to zero if the occupants are not wearing masks.

#### 2.1.4 *Breathing rate(*BR_k_)

A wide variation in breathing rate is observed in numerous studies. We have chosen to base our values on data originally reported by Ref. [43] as incorporated into the EPA Exposure Factors Handbook [44]. The estimation of breathing rate is critical to both the emission of infectious particles and exposure due to inhalation for an airborne pathogen. We have taken published tables from the handbook and adapted them to provide estimations of breathing rates for a variety of activities. With the available data, we have assumed an evenly distributed population from ages 16 to 61, with a male:female ratio of 1:1. Drawing on handbook values from Tables 6-17, 6-19, 6-40, 6-42, we created profiles for a number of different physical activities:

– sitting (office activity),
– standing (without moving),
– light intensity activity (walking, lectures, singing),
– moderate intensity activity (jogging, manual work in a laboratory or workshop),
– high intensity activity (running, exercising, heavy duty equipment manipulation, manual material transport).

The data for each activity level (Supplementary Table S.2) has been fitted to a log-normal distribution model (cf. Supplementary Fig. S.3), having one set of distribution descriptors per activity type (incorporating the variability of the population) instead of single point deterministic values.

### 2.2 Viral removal rate (vRR)

Once viruses are expelled from the exposed host, they are subject to environmental and biological effects which would reduce the viral concentration in air. The effects of air exchange, aerosol settlement, viral inactivation (biological decay) and filtering through an air cleaning systems may be considered in a simplified form by combining the contributions from these four effects into one property *λ*_vRR_ [10], representing the viral removal rate per hour, by means of the summation:

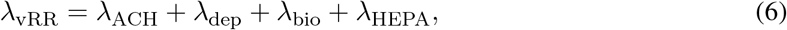

where *λ*_ACH_, *λ*_dep_, *λ*_bio_ and *λ*_HEPA_ (all in h^-1^) are the removal rates related to ventilation, gravitational settlement, biological decay and particulate filtration, respectively.

#### 2.2.1 Effect of ventilation

Effective ventilation is a known preventive measure to mitigate airborne transmission [1]. The supply of clean outdoor air, referred to as ‘fresh air’, is important to locally dilute the airborne virus and remove the pathogens by exchanging them with virus-free air.

The removal rate due to ventilation (*λ*_ACH_) via mechanical or natural means, is obtained from the amount of fresh air supplied to the space and the volume of the room:

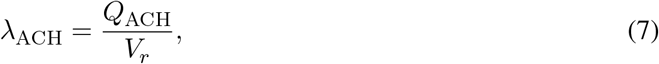

in which *Q*_ACH_ represents the volumetric flow rate of fresh air supplied to the room (in m^3^ h^-1^) and *V* its volume (in m^3^). *Q*_ACH_ will depend on the type of ventilation used.

Mechanical ventilation is considered when the indoor space benefits from active means to supply fresh air, powered by equipment such as motor-driven fans and blowers normally installed in air handling units (AHU) of Heating, Ventilation and Air Conditioning (HVAC) systems. The fresh air flow rate for mechanical ventilation is considered at the level of the supply grilles or diffusers. For energy efficiency reasons, some air handling units may be equipped with a mixing chamber to recycle part of the return air extracted from the indoor space. In this specific case, it is proposed to evaluate *Q*_ACH_ by including only the portion of fresh air supplied in the space (i.e. total supply flow of the AHU minus the recycled air flow). If the AHU is fitted with an HEPA filter, the portion of recycled air shall be included in *λ*_HEPA_ (cf. Section 2.2.4).

Although the use of mechanical systems in Europe is increasing, the greatest share of ventilation systems employ natural ventilation [45], which generates a flow of fresh air coming directly from outdoors, created by a pressure differential through permanent or temporary openings in the building’s facade. This pressure differential is caused either by: i) outdoor and indoor temperature difference, where the buoyancy force arising from gravity and the difference in air densities can be used to drive the flow, or ii) wind contouring the building structure, where the velocity profile, on both facades, creates a windward and leeward exposure. To establish a wind-driven flow, the indoor space in question shall have openings on opposite facades (windward and leeward exposure). In addition, the pressure difference depends on the mean wind boundary velocity, which fluctuates during the course of the day, ranging in intensity and geographical direction. With this said, and in view of simplifying the model, this paper will only consider a buoyancy-driven flow arising from natural ventilation.

To streamline the estimation of *Q*_ACH_, additional simplifications and assumptions are proposed. We consider single-sided natural ventilation, i.e. openings on one facade, although in reality occupants generally open windows and doors connecting to corridors or other volumes (i.e. cross ventilation). The later form of natural ventilation might extend the pressure gradient beyond the volume of the room yielding potentially higher flow rates that would reduce the risk, hence our choice is conservative. The limiting depth for effective single-sided ventilation is typically 5.5 m or up to 2.5 times the room height [46], therefore this limitation is a boundary condition for model validity.

The fresh air flow *Q*_ACH_ for single-sided natural ventilation is derived from a combination of Bernoulli’s equation and the ideal gas law [47]:

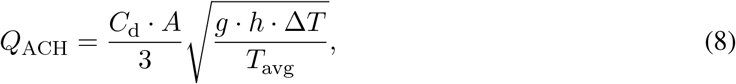

where *C*_d_ is the discharge coefficient; *A* is the area of the opening (in m^2^); *g* is the gravitational acceleration (in m s^-2^); *h* is the height of the opening (in m); Δ*T* is the indoor/outdoor temperature difference and *T*_avg_ is the average indoor/outdoor air temperature (in K). Equation (8) is valid when Δ*T* is positive and not too large (≤20 K). The CARA model incorporates a large meteorological data set of averaged hourly temperature, by month and location. These temperature profiles are based upon historical data from the HADISD.3.1.1 [48] data set. The hourly averages are computed from the past 20 years for each weather station with complete and valid data within this time range. For simplification, in this study we consider only data for Geneva, Switzerland during the months of June and December (replicating summer and winter conditions, Supplementary Fig S.4).

The discharge coefficient *C*_d_ represents the fraction of the opening area that is effectively used by the flow - it is smaller than the actual area of the opening because of e.g. viscous losses [47]. For sliding or side-hung windows, *C*_d_ is estimated at 0.6 [46, 47]. For top- or bottom-hung windows, *C*_d_ depends on the opening angle *ϕ* (in deg) and the ratio 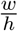 (with *w* the width of the window), according to the following rule [46]:

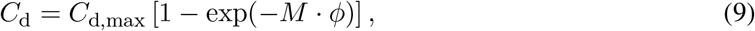

where *M* and *C*_d,max_ are given for different values of 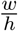 in Supplementary Table S.3. The opening angle *ϕ* can be obtained via: 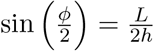, with *L* the size of the opening (i.e. such that *A* = *h · L*).

In this paper, we consider events in the winter period with a constant temperature difference Δ*T* of 10 K and in the summer with a constant Δ*T* of 2 K. The indoor temperature is also assumed constant during the exposure. As for the openings, we considered a standard sliding-type window with a height *h* of 1.6 m and an opening length *L* of 0.6 m. Examples of other window opening types can be found in Supplementary Fig.S.5. In the absence of natural and mechanical ventilation, the removal rate *λ*_ACH_ will be governed by the air infiltration of typical buildings. In this study we assume a constant average value of 0.25 h^-1^ [49].

#### 2.2.2 Biological decay

The environmental conditions have an impact on the stability and viability of the virus in air. The half life of SARS-CoV-2 in aerosols was initially measured with a median of 1.1 hours, equivalent to SARS-CoV [50]. However, in this reference the measurements were performed at room temperature (23°C) and with a relative humidity (RH) of 65%, which is not the nominal humidity level one would assume for indoor spaces in specific seasons of the year, e.g. during the winter period. The humidity of the air in the room plays a decisive role in the capacity of the viruses to survive [51–53]. Air with a low relative humidity (typically under 40%) allows smaller particles to desiccate quickly and, while the water is completely removed, the salt content of the droplet nuclei might crystallize, ending up preserving the viruses by forming a sort of protection cover. This mechanism explains why flu epidemics frequently occur during the winter period with the effect of central heating, which desiccates the air by adding sensible heat and increasing its enthalpy at a constant specific humidity. It is also apparent that the ambient humidity may play a role in the effectiveness of the bodies natural defense mechanisms against airborne viruses, with low humidity increasing susceptibility to infection [53].

However, a comprehensive model of the interplay between temperature, humidity and viral infectiousness remains a topic for further study. In this paper, we do not consider the effect of temperature on half life, although we acknowledge that there is significant data linking increases in temperature with a reported decrease in half life. We apply a simplification of the three-regime model proposed by Yang et al. [52] for seasonal influenza, with only two humidity regimes considered (RH < 40% and RH > 40%). In each humidity regime, we use values consistent with Figure 3A in [51] for the extrapolation of the half life of the virus. In the low humidity regime (RH < 40%, 20°C), virus viability after 1 hour is 70%, compared to 20% in the high humidity regime (RH > 40%, 20°C). We use the ratio of these two values to scale the half life for the low humidity regime as follows:

– In the mid/high humidity regime, we consider a half life of 1.1, based directly on Ref. [50], such that 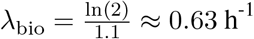
– In the low humidity regime, we apply the extrapolation based on the data from Ref. [52], and obtain a half life of 3.8 such that *λ*_bio_ *≈* 0.18 h^-1^.

The ratio of the half lives in our low humidity and high humidity regimes is 3.5, similar to that reported in the literature, notably in Ref. [54], where median half lives of 6.4 hours (RH = 40% at 22°C) and 2.4 hours (RH = 65% at 22°C) are reported in Figure 1b, giving a ratio of 2.6. The method employed in the aforementioned study, namely capture on polypropylene surfaces, is consistent with reports of longer viral half lives. We therefore consider this to be a conservative figure for the regime RH < 40%, since the reference is taken at the start of the low humidity region.

**Figure 1:**
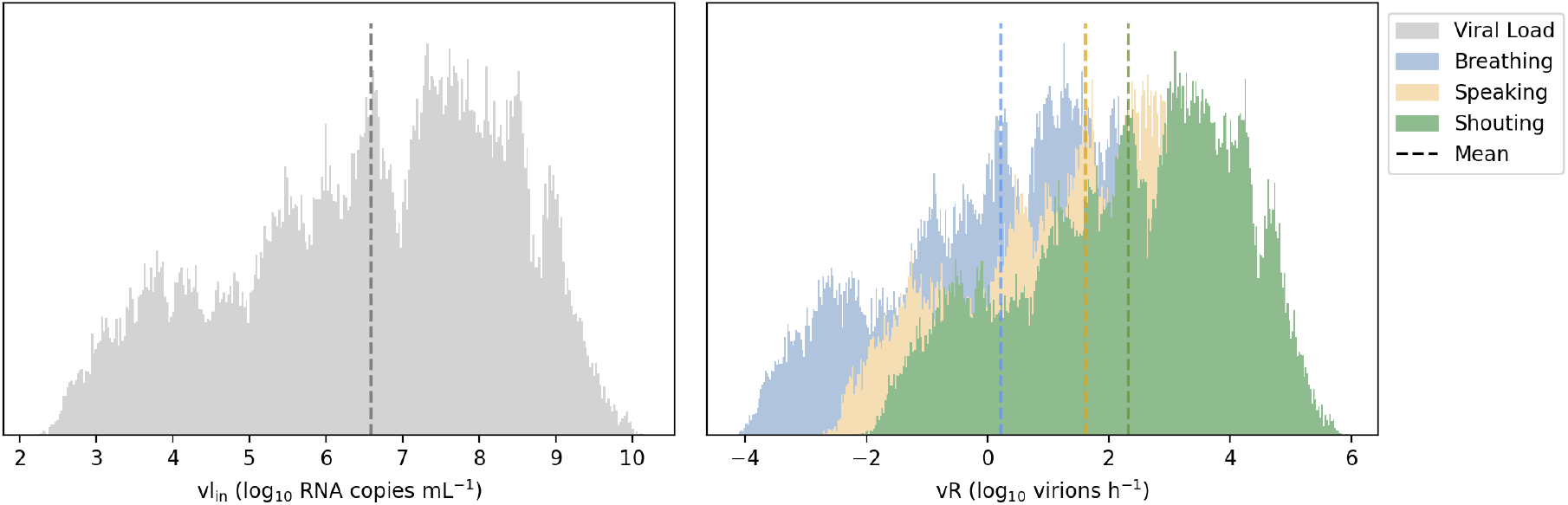
Results of the MCS of the viral emission rate distribution for an infected host breathing, speaking and shouting, while undertaking sedentary physical activity (*seated*) and comparing with the viral load distribution. Dashed lines correspond to the mean of the log_10_ values. The vertical axis of the histograms corresponds to the estimation of distribution PDFs. Median vR^total^ values: 3, 67 and 349 virion h^-1^ for breathing, speaking and shouting, respectively. The values are without the effect of face covering (*η*_out_ = 0).

In the model, we differentiate between the two regimes on the basis of the corresponding average seasonal indoor humidity, due to the effect of central heating. Unless specified otherwise, the default humidity regime discussed in Section 4 is RH > 40%.

#### 2.2.3 Gravitational settlement

Once particles are airborne, they are subject to aerodynamic forces which tend to balance with the force of gravity (dead weight), with the absence of additional momentum. Using the Stokes law, one can analytically calculate the settling velocity of a certain particle, corresponding to equilibrium between the sum of the drag and buoyancy forces and the downward force due to gravity:

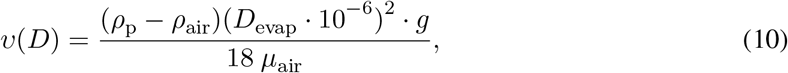

where, *ρ*_p_ and *ρ*_air_ (in kg m^-3^) are the mass densities of the particle and air, respectively; *g* is the gravitational acceleration (in m s^-2^); *D*_evap_ is the diameter (in μm) of the desiccated particle, following evaporation (*D*_evap_ = *D f*_evap_ with *f*_evap_ = 0.3), and *μ*_air_ *≈* 1.8 10^−5^ kg m^-1^ s^-1^ is the dynamic viscosity of air (at room temperature and atmospheric pressure).

Assuming the composition of a given airborne particle is dominated by water and/or organic solutes of similar density, the proposed mass density for *ρ*_p_ is 1000 kg m^-3^ [15]. The mass density of air (*ρ*_air_) is taken at 1.2 kg m^-3^. Assuming that the droplets are falling from the mouth or nose of a person standing, the height at which the terminal velocity (obtained from Eq. (10)) is reached, is considered at approximately *h* = 1.5 m from the floor, which yields *λ*_dep_ = *v/h*. Thus, particles with a diameter *D≤* 3 μm have a settling time higher than one hour and a half (and it even reaches more than 13 hours for *D* = 1 μm), whereas larger droplets with *D ≥* 10 μm are only able to maintain airborne for approximately 8 minutes and less (cf. Supplementary Fig. S.6). Therefore, the phenomenon of particle evaporation introduced in Section 2.1.2, which reduces the particles’ size, has a significant effect on the total amount of airborne particles which could potentially be inhaled by exposed hosts.

As visible from Eq. (10), the setting velocity *υ*(*D*), and hence *λ*_vRR_, depends on the diameter of the emitted particle *D*. The concentration in the air of particles of different diameters evolves therefore differently with time (see Section 2.3). Using plain Monte Carlo sampling algorithms for the particle diameters, we can also compute for *λ*_dep_(*D*) the mean (standard deviation) values, obtaining 0.054

(0.031), 0.146 (0.208) and 0.167 (0.225) for breathing, talking and shouting, respectively (all in h^−1^). The mean values are in agreement with the range adopted in other studies [55] for particles with *D*_evap_ in between 0.7 and 2 μm.

#### 2.2.4 Air filtration

The removal of airborne particles in a closed volume can be achieved by cleaning the air using High-Efficiency Particulate Air (HEPA) fibre-based mechanical filters. HEPA filters are the most efficient mechanical filters in the submicron range, increasing the probability of capturing viral-containing droplet nuclei in the air [56, 57]. The effect of this mechanism on the removal rate is determined by the volumetric flow rate of the air passing though the filter, multiplied by its efficiency (*η*_f_) and taking into account the effectiveness of the system in reducing a certain percentage of the particle load within 20 minutes (PR_20_) (Supplementary Fig. S.7). The effect of increasing the air exchange rate of the HEPA device on the particle removal efficiency can be determined by:

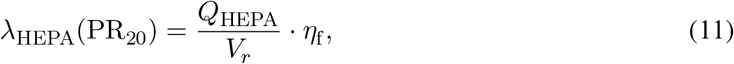

in which *Q*_HEPA_ (in m^3^ h^-1^) is the effective flow rate through the device; *V*_*r*_ is the room volume; PR_20_ is the particle removal objective; *η*_f_ is the filter efficiency. For HEPA filters certified according to EN 1822 standard [58], *η*_f_ is 99.95 % and 99.995 %, for the corresponding H13 and H14 classes, respectively. Due to the high efficiency of both filter classes, this term can be neglected. The commercial filtering device should be selected to ensure a nominal flow rate that is able to reduce sufficiently the particle load. The effectiveness of the system determines how fast the particle load is reduced in a volume. This approach is frequently used in industry, namely in the design of clean rooms [59], although this parameter is determined in the decay zone of the concentration profile 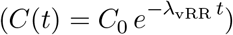 where the generating source is not present (i.e. vR = 0). In reality, the effect of a constant vR > 0 in the presence of an HEPA filter will be included in the solution of Eq. (13), therefore we use the effectiveness of the filter as an input for *λ*_HEPA_ when selecting the device. Hence, we opted to consider a particle removal objective (PR_20_), of at least 80% that would yield a clean air exchange rate *≥* 5 air changes per hour (ACH): *λ*_HEPA_(0.8) = 5 *h*^−1^, which is comparable with other design values for biological safety labs and hospital wards [60].

### 2.3 Viral concentration (*C*(*t*))

The concentration of virus depends, not only on the emission source but also on the dynamic effects linked to any potential removal mechanisms (cf. Section 2.2.1) and occupation profile (cf. Section 3), as well as possible preventive measures (e.g. face covering). This study proposes a solution of the mass-balance differential equation to simulate these effects.

The concentration of viruses in aerosols of a given size *D*, is derived from the following differential equation, determining the time evolution of the number of virions per unit volume per unit diameter, in a single-zone model:

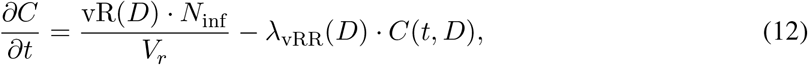

where vR has been defined in Section 2.1.2 and *λ*_vRR_ in Section 2.2, and both depends on the particle diameter *D*; *V*_*r*_ (in m^3^) is the room volume; *N*_inf_ is the number of infected hosts emitting the viruses at the same time and in equal quantities.

Solving the differential equation, we get:

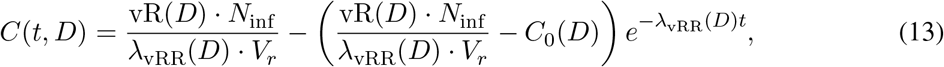

where *C*_0_(*D*) ≡ *C*(*t* = 0, *D*), and the quantity 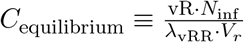 represents the equilibrium value that is reached in the steady-state regime, in which all quantities therein are diameter-dependent.

Equation (13) is valid when all variables are constant over the full time range. In our model, vR and *λ*_vRR_ may also be piecewise constant functions of time; a new value is assigned to each variable every time a condition changes in the room, in particular when an infected person(s) enters or leaves the room, or when the ventilation rates changes (which leads to a modification of *λ*_vRR_, see next Section). In-between such transition times, e.g. *t*_*n*_ and *t*_*n*+1_, all variables are constant and Eq. (13) is valid provided *C*_0_ is replaced by *C*(*t*_*n*_, *D*), and *t* by *t − t*_*n*_. *C*(*t*_*n*_, *D*) is in turn computed from the knowledge of the previous regime between *t*_*n−*1_ and *t*_*n*_; in practice all these computations are done recursively, using an efficient caching mechanism to avoid computing the same concentration twice.

### 2.4 Dose (vD)

The term ‘dose’ in this study defines the number of viable virions that will contribute to a potential infection, therefore we need to disassociate RNA copies from infectious (viable) viruses. Virus isolation from NP and throat has been largely reported, although it varies with the viral load and the number of days post symptoms onset [21, 61], which indicates a clear relation between seroconversion and viral culture, as well as the amount of RNA copies in a given sample. In addition, any existing antibodies wrapped around RNA viruses will be extracted during PCR-assayed samples. This will ‘hide’ the effect of pre-existing antibody titre in the infected host’s viral load. Hence, the proportion of virions which are viable (infectious) can be substantially lower than the measured count of RNA copies, e.g. in previously vaccinated sources [62], even though their viral loads have been reported to be similar to those in unvaccinated sources [63].

Here we estimate the receiving dose, vD (in infectious virions per unit diameter), which is inhaled by the exposed host, by first integrating the viral concentration profile (for a given particle diameter) over the exposure time and multiplying by a scaling factor to determine the proportion of virions which are infectious. Afterwards, these terms are further multiplied by the breathing (inhalation) flow rate, the fraction of viral-containing particles that deposit in the respiratory tract and the inward filtration efficiency of a face mask, if worn:

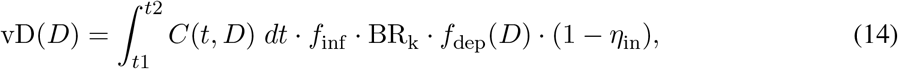

where *t*_1_ and *t*_2_ are the start and end exposure times (in h), respectively; *f*_inf_ is the fraction of infectious virus; *f*_dep_(*D*) is the (diameter-dependent) deposition fraction in the respiratory tract; and *η*_in_ is the inward efficiency of the face mask (values between 0 and 1).

Note that the breathing rate is directly proportional to the dose, hence the physical activity plays an important role in airborne transmission. Eq. (14) is valid for a single exposure from *t*_1_ to *t*_2_.

If during the simulated event, the susceptible hosts are exposed to multiple independent exposure scenarios (e.g. they leave the enclosed volume for a lunch break) or in every state-change (e.g. windows open, outdoor temperature change), the dose is given by:

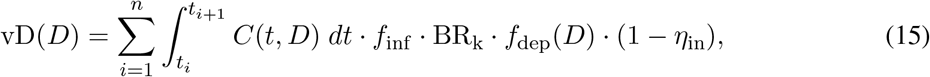

where *t*_*i*_ and *t*_*i*+1_ are the start and end times (in h) of each sub-exposure, respectively; *n* is the total amount of independent exposures in the same event (i.e. subject to the same concentration profile).

The total dose (in infectious virions) then results from the sum of all the doses accumulated for each particle size; it is given by an integral of the form

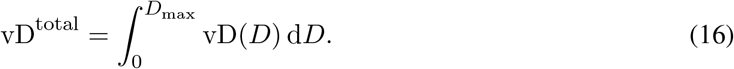

The above is computed using a Monte Carlo integration: many different diameter samples are generated using the probability distribution from Eq. (4), the dose from each of them is then computed, and their average value over all samples represents a good approximation of vD^total^ (provided the number of samples is large enough).

#### 2.4.1 *Infectious virus fraction (f*_inf_)

The aforementioned studies in the section above show the probability of isolating infectious SARS-CoV-2 viruses in serum samples increasing with viral loads larger than 10^6^ RNA copies mL^-1^, reaching a probability of approximately 90% at 10^10^ RNA copies mL^-1^ [21, 61]. For airborne samples, data also shows that only a fraction of viral RNA copies are found to give positive culture. Past studies with Influenza virus have found positive cultures in 30 % of 140 aerosol samples using a Gesundheit-II (G-II) human source bioaerosol sampler [64], corresponding to a mean swab viral load of 8.9 log_10_ RNA copies. Recent studies for SARS-CoV-2 showed successful virus isolation in 15%, 45% and 82% of samples collected from outpatients, inpatients and ICU patients, respectively [65] and 3 %, out of the 66 aerosol samples in another setting, were cultured while wearing masks, 2-3 days post symptom onset [66]. This provides evidence on the presence of infectious virus in aerosolized particles which depends on the initial viral load vl_in_, although the exact ratio between RNA copies and infectious virus is extremely difficult to determine with precision, especially when translating between *in vivo* and *in vitro* which may include other influencing factors such as post-illness onset day, the presence of antibody titre, which differ from the symptom onset, and host immunity or even the air sampling mechanism used. In absence of a simpler way of modeling these complex effects, we propose a novel approach to this parameter which can take into account the current population’s host immunity (HI), which evolves over time and requires a regular update. The higher the host immunity of the infected person, the larger is the expected antibody titre gained either from natural exposure or vaccines. The fraction of infectious virus can be estimated by:

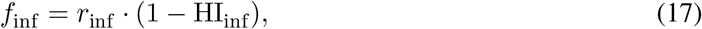

where *r*_inf_ is the viable-to-RNA virus ratio as a function of the viral load inside the infected host, with values ranging between 0 and 1; HI_inf_ represents the percentage of the infected host immunity which can be tuned to respect the up-to-date situation in any given population.

Values for *r*_inf_ are determined analysing the aforementioned data [64–66], assuming transmission in indoor settings occur up to mild/moderate illness not needing hospitalisation, we derive the following uniform distribution ranging from 0.15 to 0.45.

Increasing HI_inf_, would skew *r*_inf_ to lower values, reducing the amount of viable viruses within the count of aerosolized viral copies, since more RNA viruses are likely to be bound to antibodies. As an example, the Pfizer–BioNTech and Oxford–AstraZeneca vaccines offered a 79% and 60% protection against the S gene-positive samples (covering VOCs like Delta) [67] and, at the time of writing, 55% of the population in Geneva, Switzerland have had their second dose, exclusively from mRNA vaccine technologies. Hence, in this case: HI_inf_ = 0.79 0.55 *≈* 0.43. If HI is unknown or strapped with large uncertainties, we suggest to apply a conservative approach and assume no existing host immunity, i.e. HI = 0.

#### 2.4.2 *Inward effect of face covering (η*_in_)

In case the occupants are wearing PPE (e.g. respirators), such as N95 and FFP2, both the filtration efficiency and leak-tightness requirements are defined in the concerned test standards in the USA or European Union, i.e. NIOSH-42 CFR Part 84 [68] and EN 149 [69], respectively. Both standards use a mean particle size of a factor 10 smaller compared to those of surgical masks: 0.3 μm. According to EN 149, the material filtration efficiency and inward leakage requirements for FFP2 are 94% and 8%, respectively, providing an overall inward efficiency *η*_in_ of 87 (*±* 5) %. Despite knowing that source control measures, e.g. surgical masks, are not meant as PPE, they are still found to have an inward efficiency between 30% and 80% [41, 70]. Other studies suggest values in between 25% and 75% [71]. This variability might be linked to how well the mask is fitted to the wearer’s face. Similar to Section 2.1.3, we profit from empirical data [38, 41, 42, 71] to derive the inward efficiency of surgical masks and use the standard certification values for PPE, which include fitting requirements. Hence we propose to model *η*_in_ with uniform distributions having the following ranges of variation:

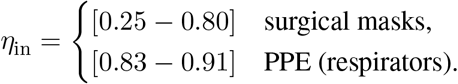

In the proposed model, a constant value of *η*_in_ is equally applied to all susceptible hosts; *η*_in_ = 0 if no masks are worn.

#### 2.4.3 *Effect of particle deposition in respiratory tract (f*_dep_)

From a pure physical point of view, the respiratory tract acts as a filter where particle deposition is distributed along its depth [27]. Similar to a mechanical filter, the three main mechanisms are i) inertial impact: large particles (*>* 2.5 μm) generally deposit in the nasopharyngeal region down to the bronchi; ii) diffusion: very small particles (*<* 0.3 μm) diffuse and deposit randomly on the surfaces of the airways and iii) sedimentation: intermediate-size particles (between 0.3 μm and 2.5 μm) - that are small enough to go into bronchioles and alveoli but big enough to avoid the Brownian motion effect - penetrate deep into the lower respiratory tract [72]. COVID-19 infections can occur from SARS-CoV-2 virus binding to ACE2 receptors which are abundant in nasal and bronchial epithelium and alveolar epithelial cells, covering both upper and lower bounds of the tract [73]. The virus can start replicating in the nose/mouth, migrating down to the airways and entering the alveolar region of the lungs to induce acute respiratory distress [14]. Therefore, it is not prudent to consider that only the smallest particles that reach the lungs contribute to the infection and we assume the total deposition in the respiratory tract, independently on the precise location. With this said, one can conclude that the fraction of inhaled particles that are absorbed in the respiratory tract (*f*_dep_) is greater than zero, however, the respiratory tract does not absorb all the infectious aerosols which are inhaled. Even if all the particles penetrate into the pulmonary region, not all are absorbed by a susceptible host since a fraction of these particles will be re-ejected once again from the airways, while exhaling, therefore 0 < *f*_dep_ < 1. A suitable practical illustration is the observation that individuals can inhale and exhale smoke particles.

In this paper, we use the well established aerosol deposition model by Hinds [72](Supplementary Eq S.1 & Fig. S.8), which is based on data from the International Commission on Radiological Protection (ICRP), averaged for males and females and at different physical activities (seated, light and heavy). In this model, *f*_dep_ depends on the aerosol particle diameter (after evaporation). One can also compute average values using plain Monte Carlo sampling for the diameter (with the BLO model introduced in Section 2.1.2), thus obtaining a mean (SD) of 0.33, (0.116), 0.484 (0.224) and 0.519 (0.228) for breathing, talking and shouting, respectively.

### 2.5 Estimation of the probability of airborne transmission

As discussed above and at the time of writing, a dose-response model for SARS-CoV-2 has yet to be developed. In this study we’ve adopted the findings by Watanabe et al. [18] for the SARS-CoV virus, where an exponential fit to a dose-response for SARS has been derived, and most recently a similar approach was adopted for a SARS-CoV-2 exposure to primates [19]. Assuming a human dose-response for SARS-CoV-2 also fits an exponential model, the probability of a COVID-19 infection is represented by

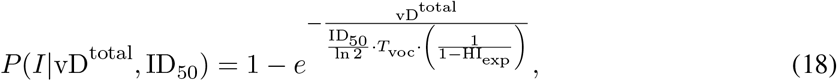

where *P* (*I*|vD^total^, ID_50_) denotes the conditional probability of event *I* (infection) for given values of the absorbed and infection doses vD^total^ and ID_50_, respectively. *T*_voc_ is the reported increase of transmissibility of a VOC, given by the ratio of basic reproductions numbers (*R*_0_) between non-VOC strains and the VOC itself (Table 2). HI_exp_ is the host immunity of the exposed occupants. The infectious dose ID_50_ corresponds to a dose required to cause infection in 50% of those exposed, and the constant ln(2) ensures that setting vD = ID_50_ (with *T*_voc_ = 1 and HI_exp_ = 0) yields a probability of 50%. Note that the inhaled viruses may find a source of resistance caused by either a pre-existing immunological condition, due to past exposure (natural or vaccine), or due to environmental conditions, such as indoor air humidity [53], which would decrease the probability of transmission for the same absorbed dose. Such effect can be considered in HI_exp_ which effectively shifts the *P* (*I*|vD^total^) curve to the right. For a hypothetical 100% immunity (HI_exp_ = 1), the infectious dose will tend to infinity. Similar to Section 2.4.1, the authors recommend a conservative approach in case the host immunity parameter is unknown, i.e. HI_exp_ = 0. On the other hand, the presence of new emerging VOCs are found to result in increased transmissibility in the population [74], which would, in turn, increase the probability of infection for a given dose. The probability of infection *P* (*I*) can be determined by integrating Eq. (18):

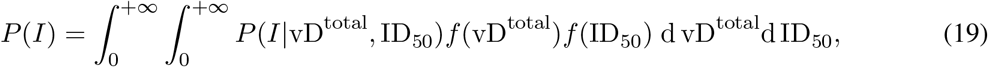

in which *f* (vD^total^) and *f* (ID50) represent respectively the Probability Density Function (PDF) of vD^total^ and ID_50_. The variability of the simulated dose is accounted for by means of a probabilistic approach using Monte Carlo sampling of the model variables (Section 2.5.2). By neglecting the effects of such a variability, *P* (*I*) would be underestimated [6]. *P* (*I*) can also be considered as the *attack rate*, where the number of new infections (*N*) is the product of *P* (*I*) by the number of exposed individuals in the room which, in turn, is equivalent to *R*_0_, if only a single individual (*I* = 1) is infected during transmission.

**Table 2:** Recommended values for *T*_voc_ based on reported increase in transmissibility [74].

| Variants | $T_{\text{voc}}$ |
| --- | --- |
| *Original strain* | 1 |
| Alpha | 0.78 |
| Beta | 0.80 |
| Gamma | 0.72 |
| Delta | 0.51 |

The exponential term of Eq. (18) considers a constant emission rate with a homogeneous mixture and a steady-state viral concentration, which varies with the ventilation rate. This study will cover the transient effects of the evolution of concentration over time, since a steady-state assumption is a limitation compared to the dynamics of real-world indoor outbreaks. Another limitation relates to the deterministic implementation of exponential relations, such as the Wells-Riley model, which is adapted to a well-known pathogen and large populations [6]. For volume-specific risk assessments, where a small population size is foreseen, a probabilistic approach is necessary.

The ratio between the number of new infections and susceptible hosts corresponds to the attack rate (i.e. infection probability) of a certain hypothetical outbreak.

The infection model only predicts transmission of secondary cases with the assumption that the incubation period is longer than the time scale of the simulation. Since the incubation period of COVID-19 is 1-2 weeks [75], the evaluation should be within this timeframe. This assumption is acceptable since He et al. [3] which found less than 0.1% of transmission to secondary cases 7 days prior to symptom onset. It is important to note that the results of infection probabilities only take into consideration the airborne transmission of the virus. It does not include short-range aerosol exposure (where the physical distance of 1-2 meters plays a critical role), nor the other known modes of transmission such as fomites. Hence, the results from this study are only valid when the other recommended public health & safety instructions are observed, such as adequate physical distancing, good hand hygiene and other infection prevention measures.

#### 2.5.1 *Infectious Dose (*ID_50_)

The number of infectious viral particles needed to cause an infection of a disease defines the infectious dose of the pathogen. Such a dose depends on various factors, such as; the type of exposure (aerosol; intranasal; fomite) causing infection and how the immune response reacts once exposed [76]. In virology, the infectious dose is normally defined using a dose-response model expressed as ID_50_ (median dose) that causes infection in 50% of the exposed individuals, *in vivo*; or, *in vitro*, when inoculating cell culture, expressed as TCID_50_ (median tissue culture dose). The precise dose-response for human hosts via airborne transmission of COVID-19 is not yet determined, hence we have opted for data reported for other coronaviruses [18]. Based on dose-response measurements of other known coronaviruses, e.g. SARS-CoV, the ID_50_ via airborne transmission was modeled at the equivalent of 280 plaque-forming units (PFU) (95% Confidence Interval (CI) from 130 to 530 PFU) [18]. Even lower values were found for other respiratory viruses like influenza, with an inhalation of TCID_50_ between 0.7 and 3 PFU that was enough to cause seroconversion, as well as prolonged wheezing and vomiting [77]. For SARS-CoV-2, a more recent study with nonhuman primates showed an exposure of 52 TCID_50_ was enough to cause seroconversion and 256 TCID_50_ for the presence of fever [19], with a dose-response curve likely to equally fit an exponential model.

Based on a preliminary collection of experimental studies and modeling estimates, the median infectious dose for SARS-CoV-2 is likely to be between 10 and 1000 infectious virions [78]. Nonetheless, in the absence of relevant statistical data, we have opted to use a probabilistic approach with values ranging from 10 - 100 infectious virions, which the authors deem reasonable for a novel agent in a fully susceptible population. This might also be a safe assumption without knowing the heterogeneous infectivity distribution of the respiratory tract such as for the influenza virus, for example, where ID_50_ is about two orders of magnitude higher by intranasal inoculation compared to aerosol inhalation [20], hence a range of values is acceptable in the absence of such data. The infectious dose is considered as a constant parameter in the model.

#### 2.5.2 *Probabilistic approach to the estimation of* vD^total^

In this paper, the dose vD^total^ is calculated solving the integrals in Eqs. (15) and (16) for some given time intervals, plugging-in the concentration function from Eq. (13), where the presented variables (e.g. *f*_dep_, BR_k_, etc.) are considered as time invariant. To account for the aleatory uncertainties, most variables are treated as random, with the result that vR, calculated by Eq. (1), is considered random as well. Concerning the viral load vl_in_, a distribution function has been obtained using the Kernel Density Estimation (KDE) technique. Table 3 summarizes the adopted distribution models and the related statistics.

**Table 3:** Summary of random variables for the infection model

| Random (stochastic) variables |  |  |  |  |  |  |
| --- | --- | --- | --- | --- | --- | --- |
| Expiratory Activity | Variable | Symbol | Mean or [Range] | SD | Unit | Fitting Distribution Model |
| All | Breathing flowrate | $BR_k$ | | | | |
| | Seated | $BR_{\text{se}}$ | 0.51 | 0.053 | | |
| | Standing | $BR_{\text{st}}$ | 0.57 | 0.053 | | |
| | Light activity | $BR_l$ | 1.24 | 0.12 | $\text{m}^3 \text{h}^{-1}$ | Log-Normal |
| | Moderate activity | $BR_m$ | 1.77 | 0.34 | | |
| | Heavy activity | $BR_h$ | 3.28 | 0.72 | | |
| | Viral load | $vl_{\text{in}}$ | 6.6 | 1.7 | $\log_{10} \text{RNA copies mL}^{-1}$ | Gaussian Kernel Density Estimation from dataset [23] |
| | Mask efficiency | $\eta_{\text{in,surgical}}$<br>$\eta_{\text{in,PPE}}$ | [0.25 - 0.80]<br>[0.83 - 0.91] | - | - | Uniform |
| | viable-to-RNA virus ratio | $r_{\text{inf}}$ | [0.15 - 0.45] | - | - | Uniform |
|  | Infectious dose | ID<sub>50</sub> | [10 - 100] | - | PFU<sup>a</sup> | Uniform |

<sup>a</sup> The dose can simply be expressed as *infectious viruses* or *viable viruses*

In the paper, we refer to *f* (vD^total^) and *F* (vD^total^) respectively as the PDF and the CDF of vD^total^. The values of such PDFs and CDFs are estimated by applying Eqs. (1), (13), (15) and (16) for each value BR_k_ and vl_in_ obtained by plain Monte Carlo Simulations (MCS) from the distribution models described in Table 3.

## 3 Occupation and activity profiles

The model supports a piecewise occupation profile where both the infected or exposed hosts can migrate in and out of the room at a given time, representing a close to real-life occupancy. In addition, we included a set of default activity profiles in terms of vocalisation activity and physical effort, which provide a weighted average of viral emissions, depending on the type of activities performed in each scenario.

The scenarios chosen in this study, and their respective baseline activities, adopted measures and geometric parameters, are summarized in Table 4. The baseline preventive measures are not a representation of any particular real-life scenario, and shall not be used as a comparison with actual settings or to local public health related measures. We consider that during the breaks, the occupants leave the room and do not gather together in another indoor space, i.e. it considers a lapse of time where the occupants are not exposed to any airborne viruses.

**Table 4:** Baseline scenarios used to generate results. By default, no masks are used. The preventive measures are not a representation of any particular real-life scenario, and shall not be used as a 1:1 comparison with actual settings or to local public health related measures

| Baseline Scenario | Activity | Occupation | Ventilation | Volume (m<sup>3</sup>) | VOC & Vaccine |
| --- | --- | --- | --- | --- | --- |
| Shared office | Office-type:<br>- Speaking 1/3 of the time, seated<br>Indoor humidity: 40 < RH < 60 % | 4 occupants;<br>1 infector:<br>- 8h workday exposure<br>- 1h lunch break | Natural:<br>1.6 x 0.2 m (partial) window opening, summer season | 50 | Delta VOC & non vaccinated occupants |
| Classroom | Training-type:<br>- Teacher: speaking, light activity<br>- Students: breathing, seated<br>Indoor RH < 40 % | 20 occupants;<br>1 infector (teacher):<br>- 8h class<br>- 1h Lunch + 2 yard breaks | Natural:<br>Periodic opening<br>1.6 x 0.2 m (partial) window opening, winter season | 160 |  |
| Ski cabin | Confined-type:<br>- Speaking, moderate activity<br>Indoor RH < 40 % | 4 occupants;<br>1 infector:<br>- 20 min ride | - | 10 |  |
| Benchmark scenario: Skagit valley chorale outbreak [79] | Singing-type:<br>- Shouting, light activity<br>Indoor 40 < RH < 60 % | 61 occupants;<br>1 infector:<br>- 2h30min exposure | Mechanical:<br>0.7 ACH | 810 | Original strain & non vaccinated occupants |
| Benchmark scenario: Bus ride outbreak [80,81] | Sedentary-type:<br>- Talking, seated<br>Indoor humidity: 40 < RH < 60 % | 68 occupants;<br>1 infector:<br>- 1h40min exposure | Infiltration:<br>1.25 ACH | 45 |  |

To benchmark our model we used a case study of the epidemiological investigation into the Skagit valley chorale outbreak by Miller at al. [79] and the investigation among bus riders in Eastern China by Shen et al. [80]. The Skagit valley chorale outbreak recorded an attack rate between 53% and 87%, and 34% in the case of the bus ride outbreak.

## 4 Results

To assess the accuracy of the model, the viral emission rate vR^total^ was benchmarked against experimental data and tuned to the datasets for SARS-CoV-2 [64, 82, 83] (Supplementary Fig. S.9), with a particle concentration of 0.06 and 0.2 cm^-3^ for the B and L modes, respectively. The mean NP(swab)-to-aerosol RNA copy number ratio (vl_in_*/*vl_out_) ranges between 2.4 *·* 10^6^ for breathing and 7.5 *·* 10^3^ for shouting.

As a result of the probabilistic approach from a MCS of 250 000 samples, the Mean (SD) of vR^total^ for breathing, speaking and shouting was 0.2 (1.7), 1.6 (1.7), 2.3 (1.7) log_10_ virion h^-1^, respectively (Fig. 1). Note that the absolute values of the emission rates differ mathematically when computing the mean: 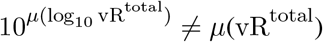. For visualization purpose we opt to show *μ*(log_10_vR^total^). The emission rate was dependent on the expiratory and physical (metabolistic) activities (Supplementary Fig. S.10). Standard vocal (speaking) activities increased the emission rate by one order of magnitude compared to tidal breathing, while louder vocalisation activities (shouting/singing) yielded an increase of two orders of magnitude. The physical activity also increased the emission rate, yet with a smaller weight compared to the activity (2.5-fold for speaking and 6.5-fold for shouting, compared to tidal breathing).

The distribution of vR^total^ ranges 7 orders of magnitude from the 1^st^ to the 99^th^ percentile. This is due to the large variability of the viral load in assessing the emission rate (Fig.1, Supplementary Fig. S.10).

Simulations where the infected host is wearing a surgical-type mask shows an average 5.3-fold reduction in the emission rate. This ratio seems to be maintained through different physical activities (5.2-, 5.4-, 5.2-fold for seated, light and heavy activities, respectively).

The successive application of Eqs. (1) and (13), generated by MCS on the distributions in Table 3, allows corresponding samples of the viral concentration *C*(*t*) (integrated over all aerosol particle diameters) to be calculated and, therefore, the estimation of its mean and significant percentiles at any given time. Following this approach for the baseline scenarios we obtain a peak mean concentration of 3 [90% CI: 1 · 10^−5^ – 10], 14 [90% CI: 5 · 10^−5^ – 47] and 220 [90% CI: 6 · 10^−4^ – 710] virion m^-3^ for the shared office, classroom and ski cabin scenarios. As for the mean cumulative dose absorbed by the exposed host (vD^total^), we obtained 3 [90% CI: 3 · 10^−5^ – 15], 10 [90% CI: 1 · 10^−4^ – 50] and 17 [90% CI 2 10^−4^ – 86] infectious viruses, for the exact same scenarios. Once again the wide confidence interval is governed by the viral load distribution, as previously discussed. We also compared the effectiveness of different measures deviating from the baseline, in order to understand their effectiveness. The results from the shared office scenario show that combining mask mandates induce a 11-fold decrease in the cumulative dose, while closing the window accounts for a 3-fold increase (Supplementary Fig S.11). In the classroom scenario, we tested different natural ventilation regimes, as well as face covering and air filtration measures. The results are plotted in Fig. 2 with source control (masks) being the most effective measure with a 12-fold decrease in cumulative dose. Closing the window or choosing to open it only during playground (yard) and lunch breaks increases the absorbed dose by a factor 2.2 and 1.6 (compared to the baseline). The effect of proper HEPA filtration is comparable with a full opening of the window during summer, between 1.6- and 2.1-fold decrease in the dose. In this study, opting to fully open a window (60 cm) in the summer is slightly more effective than partially opening the window (20 cm) in the summer, 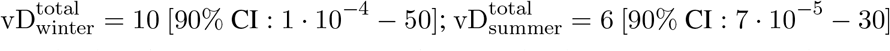 virions.

**Figure 2:**
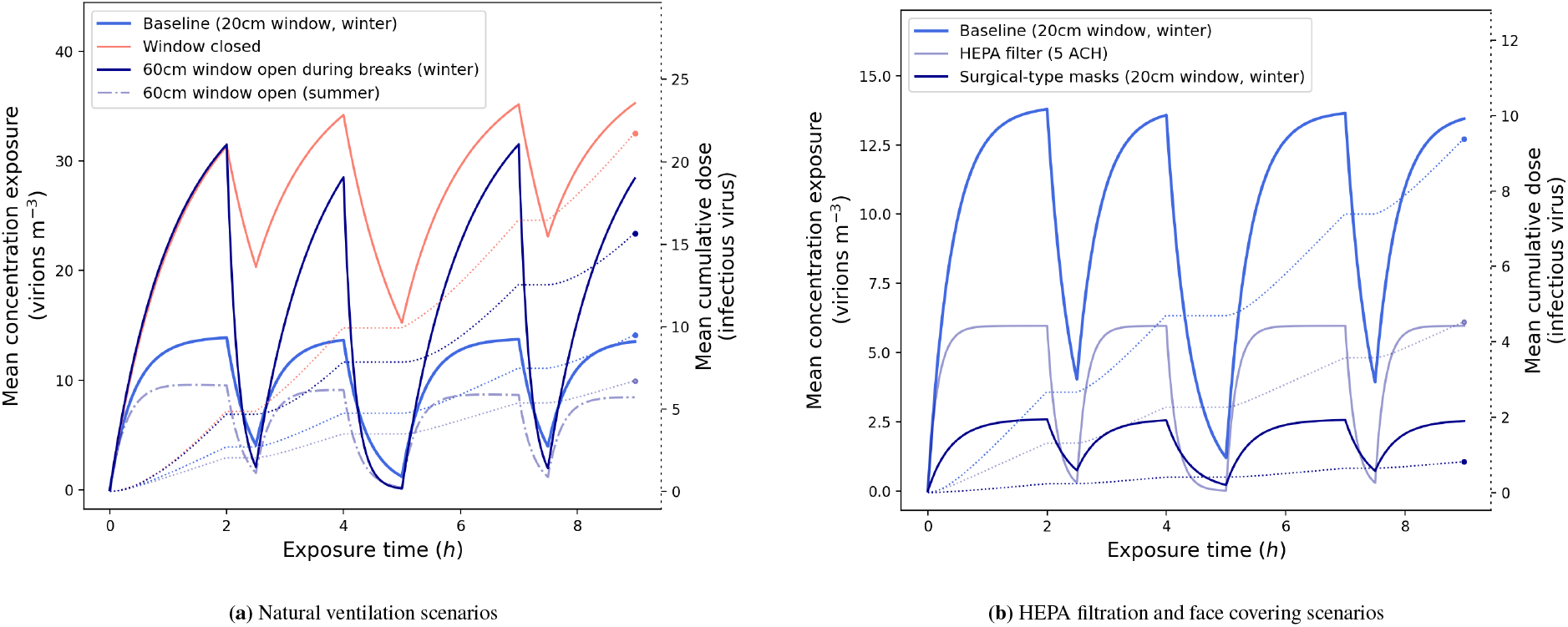
Results of the viral concentration profile over the exposure time and the cumulative absorbed dose, in the classroom scenario, for different combination of measures. The solid lines represent the concentration (left y-axis) and the dotted lines represent the cumulative dose (right y-axis). The horizontal section of the dotted lines correspond to the breaks (starting *t*=2 and *t*=7 h for 30min in the playground and *t*=4 h for 1h lunch), where the infected and exposed hosts leave the room and are not in contact for the its duration. **a)** illustrates the scenarios with different natural ventilation scenarios and **b)** the effect of HEPA filtration and masks. Note that the scale of both y-axes differ from the two subplots a) and b). For visualization purposes, the confidence interval is not represented in the figure, these values can be found in Supplementary Table S.4.

Out of the three baseline scenarios, ski cabin provides the largest cumulative dose figures, yet again yielding a 12-fold decrease in the case of the occupants wearing masks (Supplementary Fig S.12).

Solving Eqs. (18) and (19), we estimate the chances of secondary infections in each of the scenarios in the study, as well as for the two benchmark scenarios. Assuming the occupants are exposed to an equal amount of viruses performing similar physical activities, we can also estimate the number of potential secondary (new) cases, *N*, arising from an index host, by multiplying the infection probability with the *·*number of exposed hosts. The probability of disease transmission is 0.06 [90% CI: 8 · 10^−7^ – 0.37], 0.13 [90% CI: 3 · 10^−6^ – 0.78] and 0.17 [90% CI: 5 · 10^−6^ – 0.93] for the three baseline scenarios assuming the index host was infected with the Delta VOC and none of the occupants were vaccinated (Supplementary Table S.5).

Fig.3 illustrates the importance of the viral load during the potential transmission event where, e.g. in the ski cabin scenario, the wearing of masks reduces the total probability of infection (i.e. including all the random variables) from 16.9 to 3.7%. Supposing the index host is a *super-emitter*, at peak viral load of about 10^9^ copies per mL, by prescribing masks, we reduce the mean probability from 83 to 20% and can reduce even further to 6% by ensuring such rides are below 10 minutes of duration. The results without masks should be taken with caution as it may underestimate the transmission probability in this particular setting, due to potential short-range airborne exposure.

**Figure 3:**
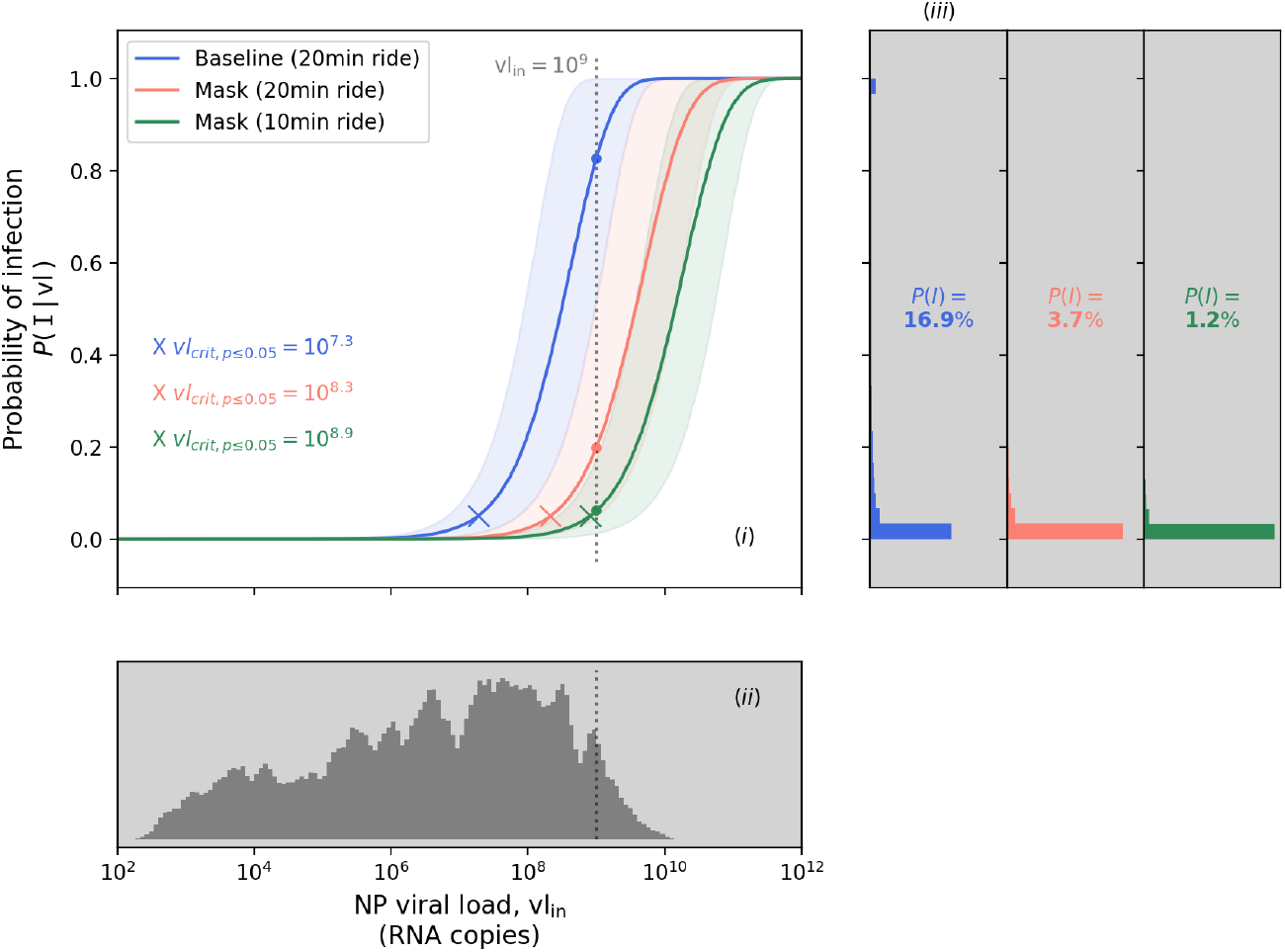
Probability of infection in the ski cabin scenario, and related dependency on the viral load. Results assume the index host was infected with the Delta VOC and none of the occupants were vaccinated. **(i)** Expected probability of infection for a given viral load value, with mean (solid line) and 90% CI (shaded area). Comparison between the baseline scenario (blue curve) and situations with stricter set of measures. The *x* markers denote the critical viral load vl_*crit,p*_ *≤* 0.05 in each situation. The dotted line correspond to the hypothetical viral load of the infected (index) host. **(ii)** Histogram of the viral load data from [23]. The vertical axis corresponds to the probability density function of the adopted distribution. The dotted line indicate the the hypothetical viral load of the infected (index) host. **(iii)** Set of histograms of the conditional probability of infection *P* (*I* vl), one for each scenario, showing the results of the MCS, including the integration on the full range of viral load data in [23]. The *P* (*I*) values shown in the middle of each histogram plot indicate the full probability (as per Eq. 19).

Our model can compare the risk with different VOCs in circulation, as well as the effect of vaccinated occupants. Fig.6 shows this comparison using as example the chorale outbreak, which had a confirmed attack rate in between 53 and 87% [79] and our model result is 72% [90% CI: 45 – 89]. If the event would have happened today, one could question what the impact of emerging VOCs and vaccination roll-out would be. Assuming the index host was infected with the Delta variant, the mean transmission probability would increase from 72 to 92% and the outbreak would have recorded 12 additional secondary infections (55 [90% CI: 41 – 59] in total). On the other hand, if all the occupants were fully vaccinated with, e.g., an mRNA vaccine offering 92 and 79% protection against S gene-negative and -positive samples, respectively [67], this would have reduced the transmission probability from 72 to 41% in the case of Delta, and to 12% in the case of Alpha. Other risk mitigation measures such as improving ventilation from 0.75 to 4 air changes per hour (ACH) are far more effective, reducing the risk from 72 to 4%.

## 5 Discussion

Modelling the emission and concentration of pollutants or harmful airborne agents in a room has been used extensively in occupational health & safety, namely for carbon dioxide and other chemicals [84]. The same fundamental physical approach was used for airborne viruses.

With this study we were able to develop a methodology and algorithm to assess the risk of airborne transmission, which can be used by facility managers, occupational health experts or interested individuals. Building on existing research done by aerosol scientists [12], we have extended the tool to include important epidemiological, virological and immunological parameters. Using this COVID Airborne Risk Assessment (CARA) modeling tool, our main findings are: 20% of infected hosts can emit approximately 2 orders of magnitude more viral-containing particles, suggesting the importance of *super-emitters* in airborne transmission; the use of surgical-type masks provide a 5-fold reduction in viral emissions; air filtration and natural ventilation through the opening of windows at all times are effective strategies to decrease the concentration of virions; slightly opening the window in the winter has approximately the same effect as a full window opening during the summer; a critical value of vl_in,0.05_ = 10^9^ RNA copies mL^-1^ could be used as the concentration threshold limit for definitions of an acceptable risk level; and pharmaceutical interventions can be included in the model to study the impact of host immunity in a given population.

Every model should be benchmarked with real-life experimental data. However, human clinical *in vivo* trial data studying the infection probability and dose response for SARS-CoV-2 is unknown to the authors. Nonetheless, we proceeded with a step-by-step validation of the model with available experimental findings and high level epidemiological investigation data for the main methods in the algorithm.

One of the major parameters in modelling airborne pollutants is the generation source term of the hazardous aerosols. Understanding this important aspect, we were cautious to analyse the model results for viral emission rate against those found in literature, across two separate domains: 1) the physiological aspects linked to the particle concentrations and size distribution and 2) the virological aspects linked to the viral load of the exhaled particles. For the former parameters, the values in Table 1 match the extensive literature review of about 20 publications by Bourouiba (Fig. 6, 7 in [33]). As for the virological aspects, the published data available is scarce [64, 82, 83], although the results of the model were still compared to studies using an human source bioaerosol sampler capable of reproducing the wanted measurements (Supplementary Fig. S.9). For breathing our model was tuned to fit experimental data on SARS-CoV-2 (measured average ratio in the order of 10^6^; model results: 2.4 10^6^), although the swab-to-aerosol viral load ratio is likely to be host-virus-specific (i.e. differing between humans and animal models) as the results in humans for influenza show higher values compared to SARS-CoV-2 (Supplementary Fig. S.9a), suggesting 1) different viruses react uniquely to the respiratory fluid properties, their location and the physical process of fluid fragmentation from mucosa to a particle spray. Since the majority of the volumetric particle emissions, while breathing come from the bronchial region, the viral abundance in that location, of the influenza patients would justify higher ratios; 2) the natural immunological control in the population, for viruses that have been in circulation for decades (such as influenza), which would correspond to higher levels of specific and cross-protective antibodies, yielding a lower amount of viable virus per RNA sample.

Comparing the emission rate results with the expiratory activities, we found one order of magnitude increase from breathing to speaking and a further order of magnitude increase from speaking to shouting, which confirms the importance of vocalisation in indoor risk assessments. In fact, the expiratory activity has a overall higher effect on viral emissions increase than the physical activity, since someone shouting while seated yields larger emission rates than breathing under heavy physical activity (e.g. while at the gym). As expected, the distribution of vR^total^ is highly governed by the range of viral load data used in the simulation, visually illustrated by the shape of the PDFs in Fig. 1. Although one could define boundaries to reduce the variability of the data, we would lose the real-life effects of the particular dynamics of this disease.

Comparing these results with the Skagit Valley Chorale superspreading event [79], where the authors used the Wells-Riley formulation to derive the so-called *quanta of infection* using reverse engineering from the outbreak investigation. The *quanta* (q) can be considered as a cluster of inhaled pathogens (SARS-CoV-2 virions in our case) required to cause infection in 63% of those exposed [85]. In other words, q can be interpreted as the number of inhaled virions divided by the infectious dose at 63% probability 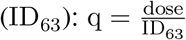, where ID_63_ = ID_50_ */* ln (2). For the chorale outbreak, a mean *quanta* was estimated at 970 q h^−1^, assuming an infectious dose between 10 and 100 infectious virions we get vR^total^ from 1.4 10^4^ to 1.4 10^5^ virion h^−1^. This would correlate for our results in the range of the 75^th^ and 95^th^ percentiles for someone shouting undergoing light activity (Supplementary Fig. S.10), which would be consistent with a superspreading scenario reported by the authors and the findings by Endo et al [86] which modelled about 10% of infectious individuals (i.e. 90^th^ per.) are responsible for the majority of secondary infections. These results could also indicate that the index host of the chorale outbreak had a viral load in between 10^8^ and 10^9^ copies mL^-1^. Such findings also support the notion of *super-emitters* in airborne transmission where a small subset of infected hosts (>80^th^ per.) emit approximately 2 orders of magnitude more viral-containing particles, compared to the median, for any given expiratory activity. In part, it may correlate to a few individuals which are found to emit much more particles than others [16] and also shed much more viruses [23]). Another outbreak during a bus ride to a worship event in eastern China, with a potential for airborne spread, was reported [80] and a quantum rate of 45 q h^-1^ ([6.5 10^2^− 6.5 10^3^] virion h^-1^) was derived based on the epidemiological study [81]. In our results, this value would correspond to an index host speaking while seated, in the 70^th^ to 90^th^ percentile range and could indicate a viral load during transmission in between 5 10^7^ and 5 10^8^ copies mL^-1^. Speaking during the voyage could well be considered as an accurate assumption since the occupants of the bus where all attending the same worship event [80], having similar interests and a common topic, leading to a sustained conversation during the duration of the bus ride. Such assumption might not be extrapolated to similar exposures in public metropolitan transpiration, where the occupants are generally not speaking to each other. Furthermore, the results of vR^total^ are also cross checked against literature data gathered by Mikszewski et al [87] (with the Wells-Riley approach), converted into virions to compare with vR^total^ (Fig. 4). Note that the literature values used in Fig. 4 were normalized to the same infection coefficient, independent on the type of virus. Knowing that viruses might differ in terms of virulence, this assumption is deemed acceptable since the original reverse engineering application of the Wells-Riley equation from epidemiological data does not include any viral infection dose [13].

**Figure 4:**
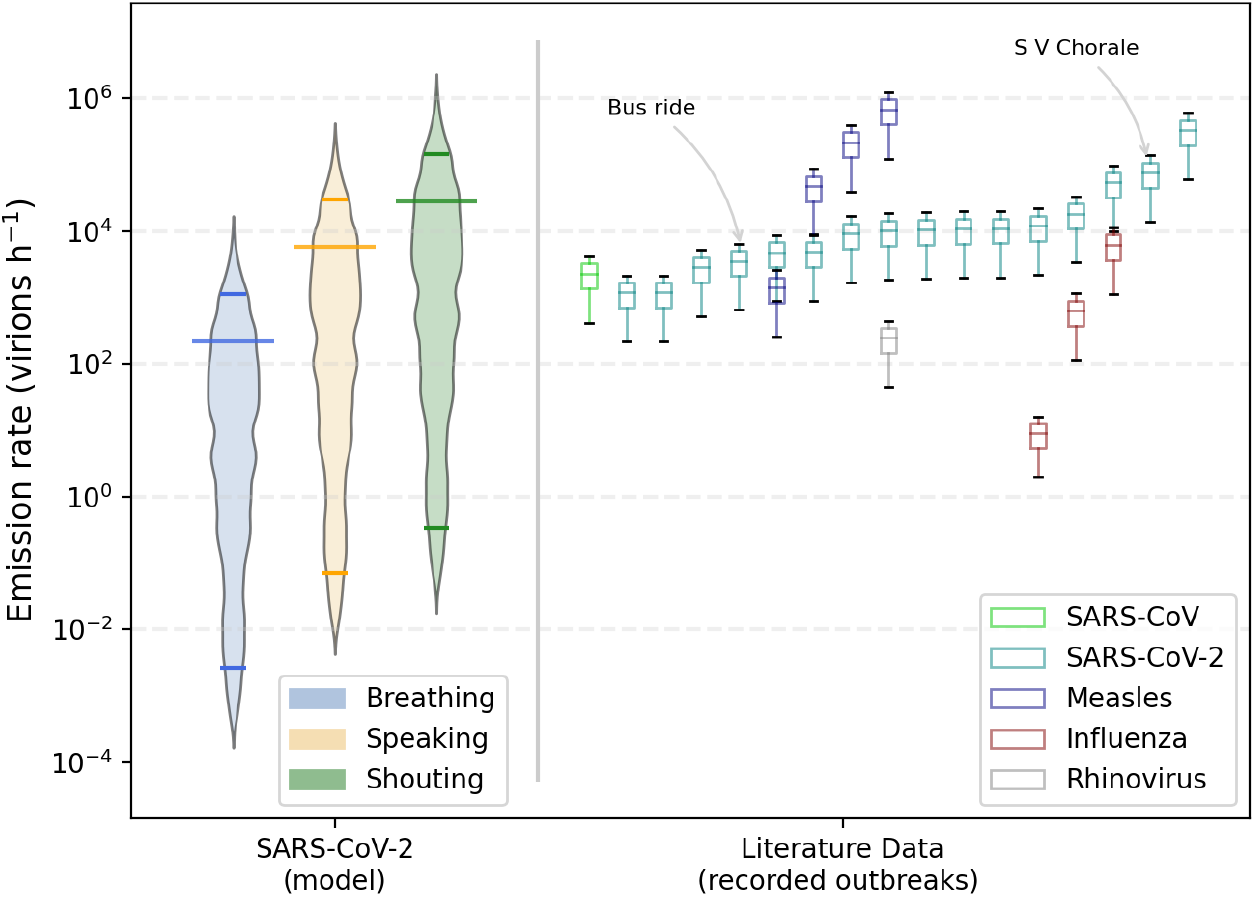
Comparison of the viral emission rate from this study with those reported in outbreaks. **SARS-CoV-2 (model)** reflects the result from the MCS for a light physical activity and different expiratory activities (Breathing, Speaking, Shouting). The violin plots denote the histograms of vR^total^, with the bottom and top bars indicating the 5^*th*^ and 95^*th*^ percentiles and the larger bar in-between indicating the mean. **Literature Data (recorded outbreaks)** is a collection of emission rate values published by Mikszewski et al [87], with values in *infectious quanta* (following the Wells-Riley approach adopted by the authors), converted into virions, normalized with an infection coefficient by multiplying 50 the values in Table 3 of Ref. [87] with the infectious dose distribution used in this study (vR^total^ = *quanta \** ID / ln(2)). The boxplots illustrate the result of this *quanta-to-virion* conversion, denoting the mean (IQR), minimum and maximum values of each distribution.

The results for the use of face masks in the model is in agreement with Asadi et al., where the effect of particle emissions with surgical masks was studied and measured a factor 6 reduction [38], compared to a 5.3 reduction factor given by our model.

We assume a homogeneous dispersion of virions in the room, hence potentially underestimating the infection risk for the occupants in close proximity to the infectious source [6]. In epidemic modeling, adopting the homogeneous mixture assumption is generally more reasonable than theoretically reconstructing the layout, airflows or interpersonal distances of the precise event where the transmission took place [20]. This assumption implies that: 1) a proper interpersonal distance of at least 1.5 − 2 m is ensured; 2) a single-zone ventilation mode; and 3) occupants are not in the same ventilation streamflow. These conditions (in particular the interpersonal distance) imply a slight overestimation of the risk of the long-range airborne transmission for short-term exposures, due to the time needed for the viruses to disperse and mix within the volume. The assumptions could be relieved by performing case-specific Computational Fluid Dynamics (CFD) simulations at the extra cost of a dramatic increase in complexity and computational time, thus hindering the benefits of a quick and easy risk assessment. Nonetheless, the authors are investigating on an analytical approach to include short-range airborne transmission in this infection model, as a potential future upgrade.

The simulations for typical shared offices show that exposure to airborne viruses is almost negligible when the occupants are wearing masks and slightly opening the windows ensuring a minimum amount of air exchange (maximum ACH: 3.8 h^-1^ in this case).

Recent guidance has emerged to encourage natural ventilation in classrooms [88], hence we focused on the different possible modalities adapted to a typical school routine. The absence of ventilation in the classroom is clearly a situation to avoid and simply fully opening the window during playground and lunch breaks (ideally to avoid the fresh air intake when the room is occupied) is not very efficient, resulting in a slight reduction (1.4-fold in exposure). Whereas, keeping a slight window opening during the winter is 60% better, yielding a 2.2-fold decrease in the dose. This suggests natural ventilation through the opening of windows at all times is an effective strategy to decrease the concentration of virions in the air, contradicting published guidance in Europe [89]. We also tested the comparison between summer and winter seasons and found that slightly opening the window in the winter has approximately the same effect as a full window opening during the summer.This result was expected since the fresh air flow for single-sided natural ventilation is proportional to 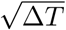 - representing the difference between outdoor and indoor temperature. This finding could be of high importance in settings where the occupants may argue against this particular measure.

In practice the use of natural ventilation via open windows might be found to be either i) uncomfortable for children due to low outdoor temperatures during the winter; ii) highly polluted outdoor air or iii) a source a distraction from external nuances. The best solution would be to equip schools with properly sized mechanical HVAC systems, although it is sometimes complicated to perform retrofitting works within existing installations. A quick, easy, affordable and effective solution would be the use of HEPA filters. Installing HEPA filters ensuring, as a minimum, *λ*_*HEPA*_(0.8) = 5 h^−1^, would reduce the mean absorbed dose by a factor of 5 compared to having the windows closed without further measures. Measurements performed in classrooms reported a similar result showing that inhaled dose is reduced by a factor of 6 when using air purifiers at 5.7 h^-1^ [90].

To summarise, natural ventilation is less effective during the summer period, although still more effective than the most conservative periodic venting scenario in Ref. [89]. Natural ventilation is, nonetheless, very important and our study would suggest leaving the windows open at all times for maximum viral removal efficiency. Analysing the effect of natural ventilation from a slightly different angle, a study has shown that higher airborne pollen concentrations might have an effect on increased infection rates [91]. Hence, opening the windows during the local pollen season may also induce a second order, detrimental effect on the infection probability which is not included in this study. However, it is safe to say that HEPA filtration will also help in reducing the pollen load in a given volume providing an extra mitigation measure against this effect. A further study could aim at including the seasonal pollen load as a variable in the model.

The model also exposes the relationship between viral load (vl_in_) and transmission probability. The conditional probability of transmitting the disease to other occupants for a given viral load value in a defined indoor environment can be evaluated by analysing 3 different zones in Fig. 5. For viral loads below a critical threshold value vl_*crit,a*_, the probability of infection is close to 0%, whereas above vl_*crit,b*_ the probability is close to 100%. This demonstrates the aforementioned importance of the viral load of the infected host at the time of transmission, showing how to maximise the chances of breaking the chain of transmission - i.e. reaching *P* (*I*|vl) *≈* 0.

**Figure 5:**
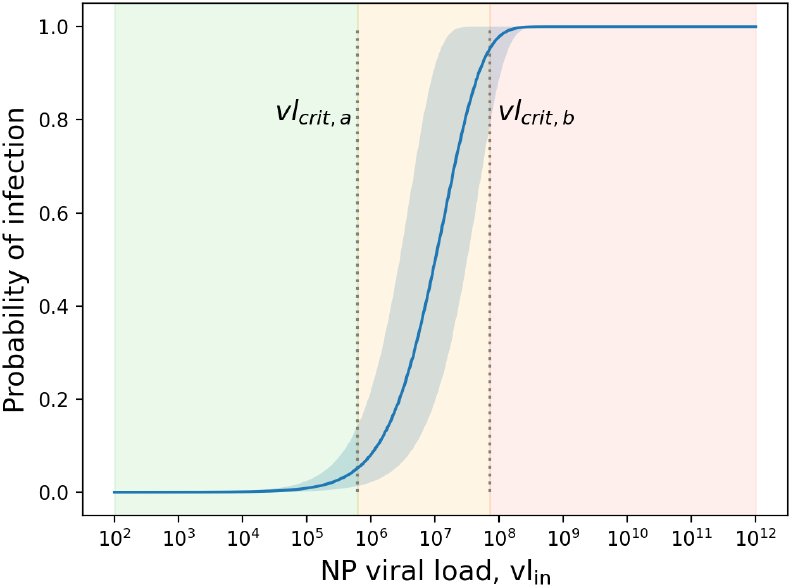
Conditional probability of infection *P* (*I*| vl), with a 90% CI (blue shaded area). vl_*crit,a*_ and vl_*crit,b*_ are the critical threshold values up to which the probability of infection is close to 0 and 1, respectively, dividing the range of viral loads into three shaded regions (in green, orange and red).

By defining the critical limits at 5 and 95%, the threshold values can be annotated as vl_*crit,p≤*0.05_ and vl_*crit,p≥*0.95_. They depend on the effectiveness of the prevention measures; by adding stricter measures the graph would move to the right, whereas relaxing the measures would shift the values to the left. Therefore, a less conservative approach in terms of preventive measures would increase the likelihood of infectious hosts with lower viral loads transmitting the disease, as shown in Fig. 3.

Some countries opted to keep ski resorts open during the widespread COVID-19 restrictions. For a typical 10 m^3^ ski lift cabin, the recommended maximum travel time is approximately 10 minutes with surgical type masks. Ski cabins generally have a small opening on one side, although if we assume that the volume is not actively heated (e.g. sensible heat from radiators) or passively heated (e.g. latent heat from the occupants), the outdoor and indoor air temperature can be assumed to be in equilibrium. Hence the effect of removal rate from natural ventilation (including infiltration) is neglected, as a conservative approach. The data also indicates that the probability of infection follows a quasi-binary relationship, i.e., for a given scenario either transmission will occur (*P≈* (*I*) 1) or will not (*P ≈* (*I*) 0). This can be observed by analysing the histograms in Fig. 3 (iii), where the majority of the samples generated by MCS on viral load distributions lead to a value of *P* (*I*|vl) in the neighborhood of the lower and upper bounds 0 and 1. The probability of falling within the orange zone in the baseline scenario (vl_*crit,p*=0.05_ *<* vl_*in*_ *<* vl_*crit,p*=0.95_) is 28%, which is spread throughout the range 0.05 *< P* (*I* | vl) *<* 0.95. In our study, the rise in probability of infection for several baseline scenarios occurs at viral loads higher than 10^6^ RNA copies mL^-1^, which strikingly correlate to the findings of van Kampen et al. [61] where the probability of isolating infectious SARS-CoV-2 viruses in RNA samples starts to increase in the same range. A deeper analysis of Fig. 3 reveals the importance of introducing appropriate measures that would shift the curves in plot (i) towards the right. Relaxing preventive measures (i.e. shifting the curve to the left) would yield higher density of samples close to *P* (*I*) *≈* 1 and therefore increase the chances of transmitting the disease. This also shows the importance and effectiveness of large scale diagnostics in asymptomatic or presymptomatic hosts early into their infection, so that they are placed in isolation before the viral load increases beyond the critical value.

By adopting appropriate measures, tailored for the specific indoor environment, the user could apply this critical threshold approach vl_*crit,p≤*0.05_ as a goal / objective for the risk assessment. Approximately 80% of the viral load samples in Ref. [23] are less than 10^8^ RNA copies mL^-1^ and approximately 95% less than 10^9^ RNA copies mL^-1^, therefore by applying control measures such that even with a potential viral load during transmission of the infected host up to *≥* 10^9^, one still ensures a significantly reduced chance of on-site transmission, i.e. vl_*crit,p≤*0.05_ 10^9^, hence the risk assessment of airborne transmission could be considered as acceptable. The residual risk linked to the remaining 5% of viral loads above *≥* 10^9^ RNA copies mL^-1^ might not be acceptable in settings which possess identified superspreading characteristics, such as Crowded, Close-contact and Confined settings - three C’s (3C), or for settings involving a large gatherings of people, such as conferences, social events or concerts. For such settings we recommend to either i) increase the threshold to vl_*crit,p≤*0.05_ 10^10^ RNA copies mL^-1^, which would probably require a non-negligible upgrade of venue layouts and ventilation systems, or 2) include, in addition, other diagnostic measures such as a rapid antigen testing strategy of the participants. The latter option would cover the residual risk of infected hosts with higher viral loads (>10^9^), where this type of diagnostic technique is most effective.

In this study, we also looked at the effect of pharmaceutical interventions, such as vaccination, and epidemiological characteristics related to emerging variants of concern (Fig.6). Our findings suggest the increase transmissibility of a given variant surpasses the counter effect of the vaccination when the protection level (against the S gene sample in question) is lower than 1 *− T*_VOC_ - in the case of the Delta VOC, a protection of 49% would be equivalent to an non-vaccinated host infected with the original (wild) strain. With a vaccine providing 79% protection against Delta, the chances of on-site transmission reduced by a factor 6-fold. This analysis is valid as long as average viral loads remain similar from vaccinated to non-vaccinated hosts; such parameter can be included in the model once accurate data is available. Since the reasons for increase transmissibility is unknown (e.g. spike mutations that would alter the infectious dose or an increased level of viral shedding, among others), a dedicated parameter such as *T*_VOC_, which is based exclusively on epidemiological data, is most accurate. Vaccination and host immunity is an important preventive and protective measure, however, according to this model, non-pharmaceutical interventions such as ventilation, seem to be much more effective with a 14-fold reduction. Hence, measures which reduce the viral density in the air should be actively supported and included early in the risk assessment process. When performing risk assessments, it is very important to adopt the hierarchical pyramid approach for protection measures, starting from eliminating, substituting or reducing the hazard, following with engineering/scientific measures and administrative controls, and leaving the prescription of PPE as a last (final) layer of protection, if needed.

**Figure 6:**
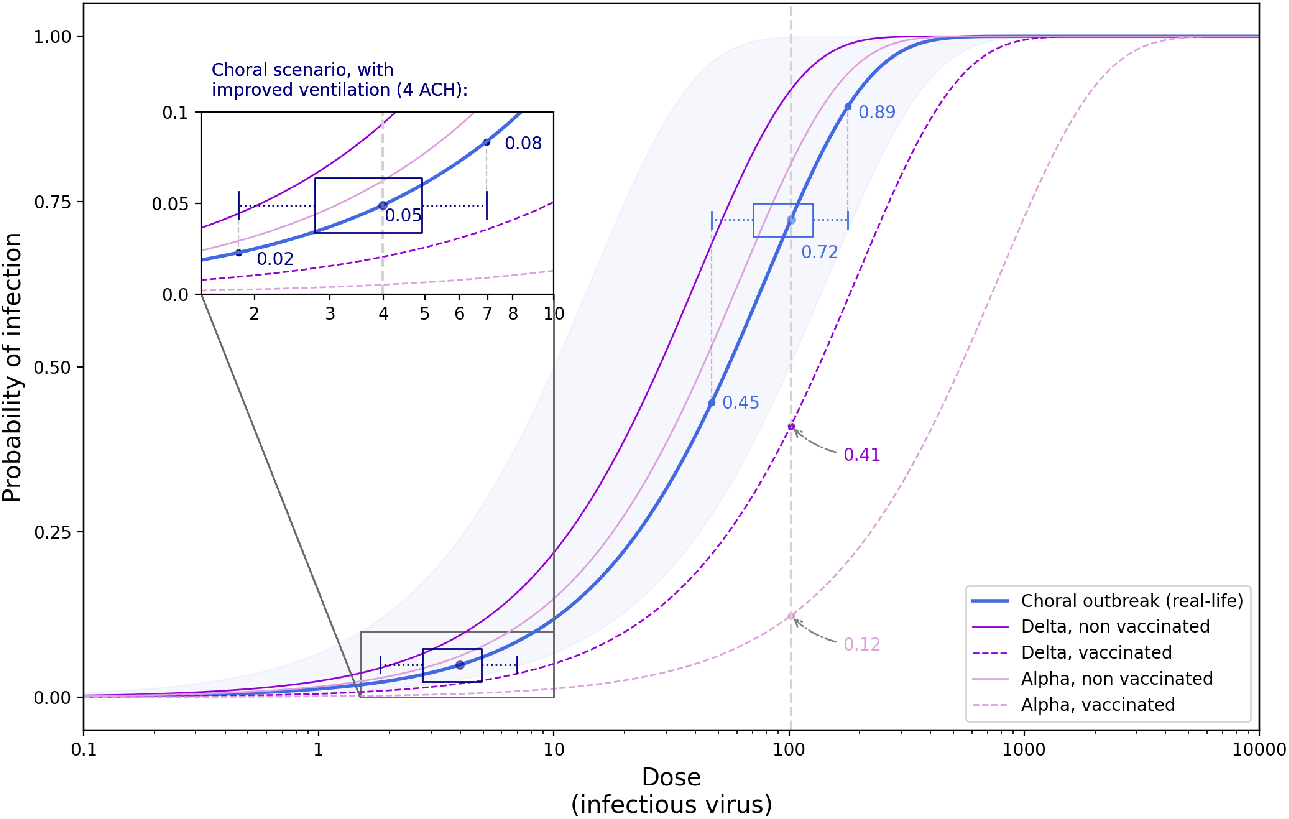
Probability of infection depending on the absorbed dose of infectious virus (vD). Comparison between the real-life outbreak scenario at the Skagit Valley Chorale event [79] and how it would relate to the epidemiological conditions 18 months into the pandemic. The solid blue line represents the model results of real-life scenario, with the shaded area corresponding to a 90% CI. The other solid / dashed lines represent the conditions with different VOCs and vaccination status. The boxplots indicate the distribution of vD for the real-life scenario and with a hypothetical ventilation improvement for comparison, with the box descriptors indicating the mean (IQR) and the whiskers indicating 5th and 95th percentile.

To maximize the benefit and global use by the public, it is important that such models are accompanied with a functional user friendly interface where the input parameters are simple and relatively easy to identify by non-experts. An example is provided for the CARA calculator tool (Supplementary Fig S.13).

## 6 Conclusions

Just like any other occupational health and safety risk, it is of vital importance to fully understand the hazards and the rationale behind the preventive measures. A proper understanding of how respiratory viruses are transmitted is an essential step towards ensuring proper protection. This paper focuses on describing the airborne transmission mode of SARS-CoV-2 and proposes a multidisciplinary approach to assess the most suitable preventive and protective measures. Facility managers, health and safety professionals, as well as individuals must systematically address the risk of airborne transmission of respiratory pathogens in indoor settings. Providing easy access to such models, despite their intrinsic complexity, accompanied by a proper user friendly interface, will greatly facilitate the required analysis to estimate the risk level. Although the notion of acceptable risk depends on national legislation and corporate/organisational risk management strategies, this paper provides some guidance on how to determine whether or not the risk of airborne transmission is mitigated. The COVID Airborne Risk Assessment (CARA) tool, allows for such a quick and accurate assessment of the indoor setting, which has been benchmarked against epidemiological and experimental data, as well as other published findings. Having a simplified model obviously relies upon some of the assumptions and consequent limitations that were discussed and justified in this study.

The present methodology is highly dependent on the viral load data and associated statistical descriptors. The use of other datasets would have an impact on the results, which might modify the findings of this study. Nonetheless, the authors were cautious to choose a distribution that would represent a broad envelope by taking data during the exponential growth phase of the epidemic, yielding conservative values compared to other less critical datasets, although this could be considered as a vulnerable aspect of the model, it is one that can be tuned once further data is published. The same can be said with respect to immunological effects. Plugging in data on the waning levels of population immunity (from either natural infection or vaccination) from the time post-infection, or post-inoculation for different vaccine technologies, would increase the accuracy of the dose-response relation. Hence, detailed modelling of host immunity dynamics, as well as short-range airborne transmission (discussed above), will be subject to a future upgrade of CARA.

To conclude, this study shows that with a risk-based approach, transmission can be mitigated in existing infrastructures without major modifications or costly consolidations plans (e.g. optimizing of the exposure time / occupation profile, ensuring sufficient natural ventilation adapted to the different temperature profiles or using adequate face covering measures). In a post-COVID era, we will face a new paradigm with the inclusion of this novel occupational hazard, using models and tools such as CARA to endorse healthy buildings and protect their occupants against respiratory infections.

## Data Availability

The CARA model and data to reproduce the results are under CERN copyright, and available on Apache 2.0 open source license from our code repository. The terms and conditions for use, reproduction, and distribution can be found under the 2.0 version of the Apache License.

https://gitlab.cern.ch/cara/publications

## Acknowledgements

We wish to thank CERN’s HSE Unit, Beams Department, Experimental Physics Department, Information Technology Department, Industry, Procurement and Knowledge Transfer Department and International Relations Sector for their support to the study. Thanks to Doris Forkel-Wirth, Benoît Delille, Walid Fadel, Olga Beltramello, Letizia Di Giulio, Evelyne Dho, Wayne Salter, Benoît Salvant and colleagues from the COVID working group for providing expert advice and extensively testing the model, as well to Alessandro Raimondo and Manuela Cirilli of the Knowledge Transfer activities.

HadISD.3.1.1 data were obtained from http://www.metoffice.gov.uk/hadobs/hadisd on 1st June 2021 and are British Crown Copyright, Met Office 2020, provided under an Open Government License.

## Authors’ contributions

AH, NM and LA conceived and prepared the study. JT supervised the work from a health science perspective. NM, PE and MR designed the mathematical model. AH, NM, LA, JD, GA, MA, PE, MR, NT developed the CARA tool used in the study. JD and PE developed the location-specific temperature profiles. AH and LA carried out the simulations. AH, LA and MR designed the figures. AH drafted the manuscript and NM, JT, LA, JD and MA revised it critically. All authors reviewed and approved the final manuscript.

## Footnotes

* The total emission rate, vR^total^ in virion h^-1^, can be obtained by integrating the emission rate over the diameter *D*.

