## Supplementary Material for "Modelling airborne transmission of SARS-CoV-2 using CARA: Risk assessment for enclosed spaces"

#### S.I Equations

The total deposition fraction, $f_{\text{dep}}$, as a function of particle diameter $D$, is, according to Hinds [1]:

$$f_{\text{dep}}(D) = I_{\text{frac}}(D)\left(0.0587 + \frac{0.911}{1+e^{4.77+1.485\cdot\ln D_{\text{evap}}}} + \frac{0.943}{1+e^{0.508-2.58\cdot\ln D_{\text{evap}}}}\right), \qquad \text{(Eq S.1)}$$

with

$$I_{\text{frac}}(D) = 1 - 0.5\left(1 - \frac{1}{1+0.00076\cdot D_{\text{evap}}^{2.8}}\right),$$

and $D_{\text{evap}} = f_{\text{evap}} \cdot D$ (with $f_{\text{evap}} = 0.3$, $D$ in $\mu$m).

#### S.II Figures

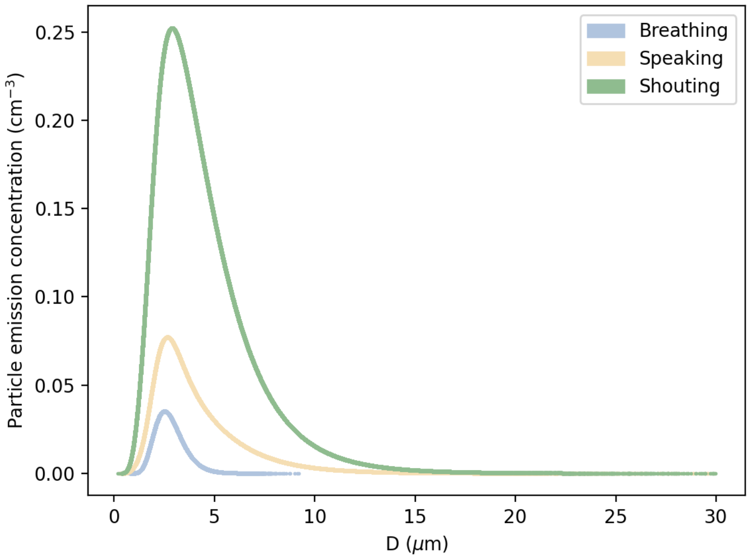


**Figure S. 1:** Particle emission concentration as a function of it's diameter.

Submitted for review

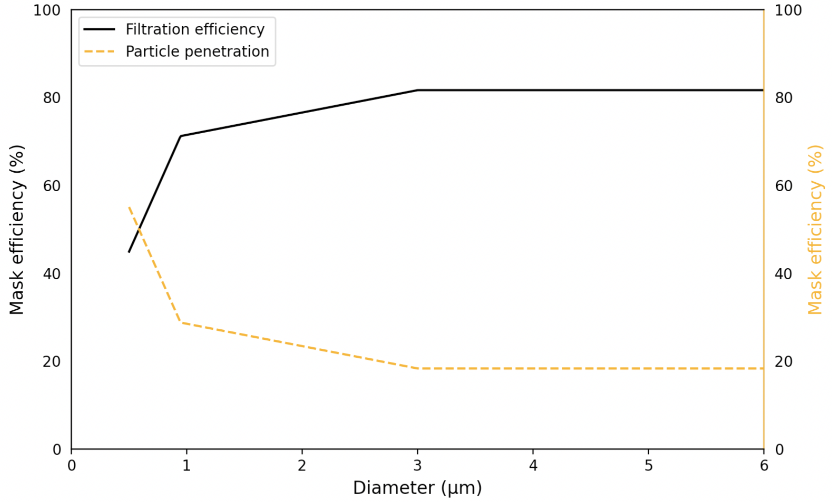


**Figure S. 2:** Effect of surgical mask in the outward direction. Black solid line represents the outward filtration efficiency of the mask, taking into account the leakages, based on empirical data; see Refs. [2–4]. The orange dashed line is the particle penetration yielding the relative fraction of particles that contribute to vR for different diameters. Note that, for D $\geq$ 3 μm, the filtration efficiency is assumed constant at 82%, while it is zero below 0.5 μm due to the lack of experimental data at the time of writing.

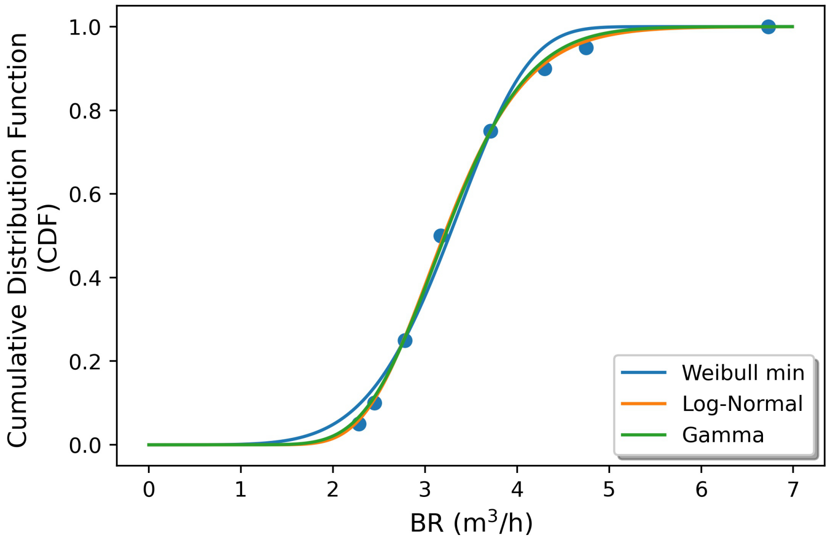


**Figure S. 3:** Outcomes of the fitting algorithm for the breathing rate distribution during a 'heavy exercise' activity, for three different kind of distribution functions, using the values from Supplementary Table S2. The best fit is obtained with a Log-Normal distribution: mean (SD) of 3.28 (0.72) $m^3\ h^{-1}$.

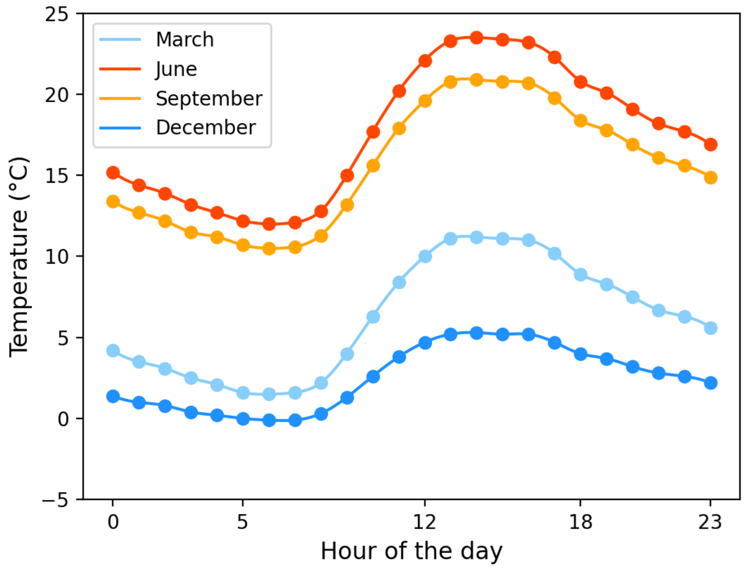


**Figure S. 4:** Average hourly temperature for Geneva, Switzerland. Data from hadISD [5].

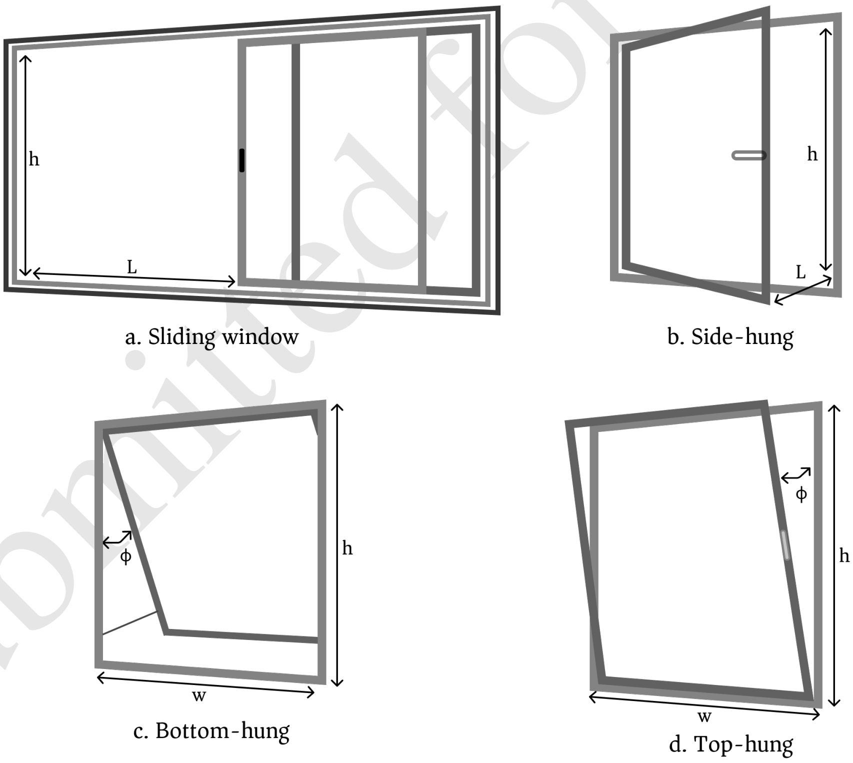


**Figure S. 5:** Example of window opening types for natural ventilation.

Submitted for review

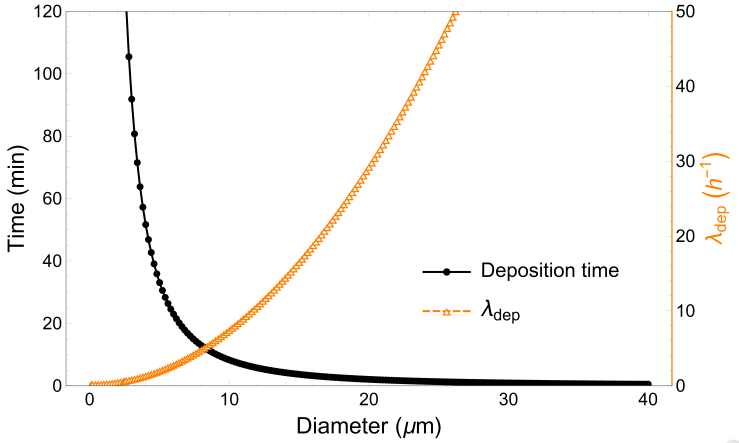


**Figure S. 6:** Effect of gravitational settling in standard indoor environments with typical air flow conditions. The solid line with circular markers represents the settling time of particles as a function of their size, assuming a terminal velocity at $h = 1.5$ m from the floor. The dashed line with the triangular markers represents the removal rate due to gravitational settling.

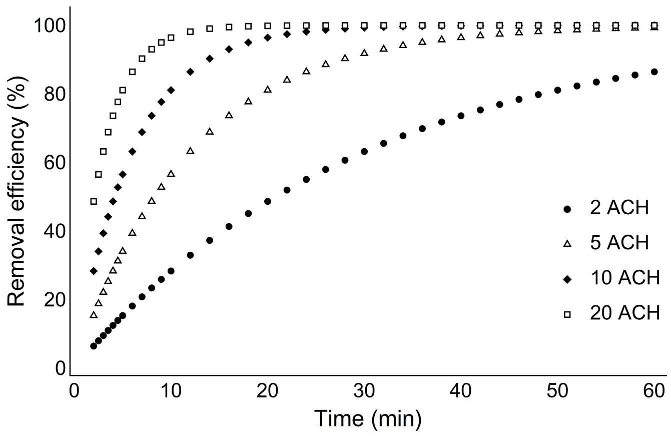


**Figure S. 7:** Time required to remove a fraction of the total particle load with HEPA filtration as a function of the mechanical performance of the device (in ACH). To reach a reasonable removal efficiency (e.g. 80 %) in an acceptable time frame (e.g. 20 min), we would need to select a device that would provide a removal rate ($\lambda_{\text{HEPA}}$) of about 5 ACH.

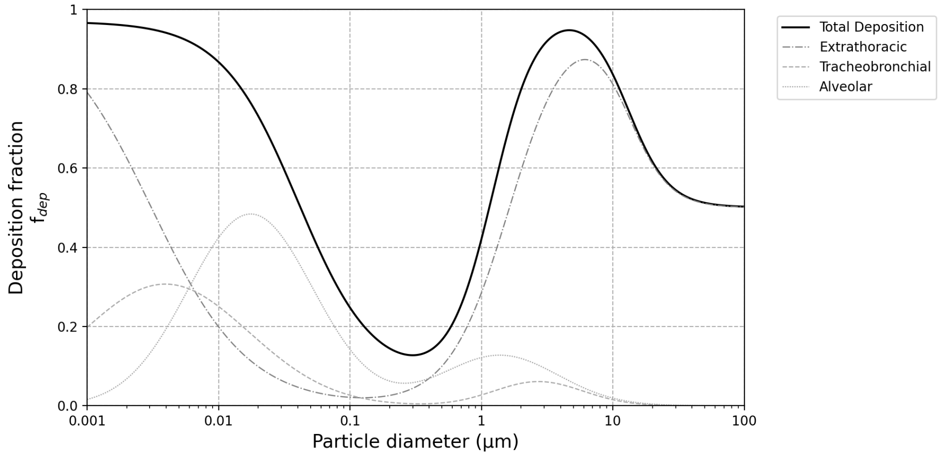


**Figure S. 8:** Representation of the ICRP deposition model published by Hinds [1], as a function of the particle diameter.

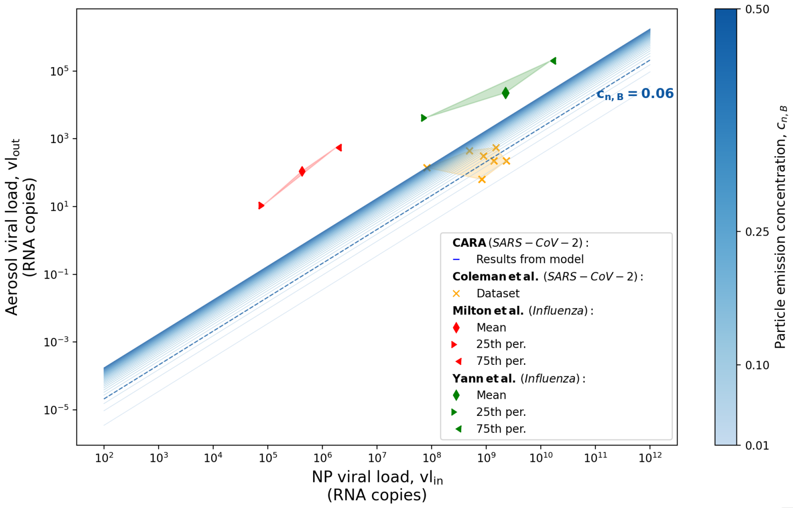


**(a)** Breathing for 30 minutes

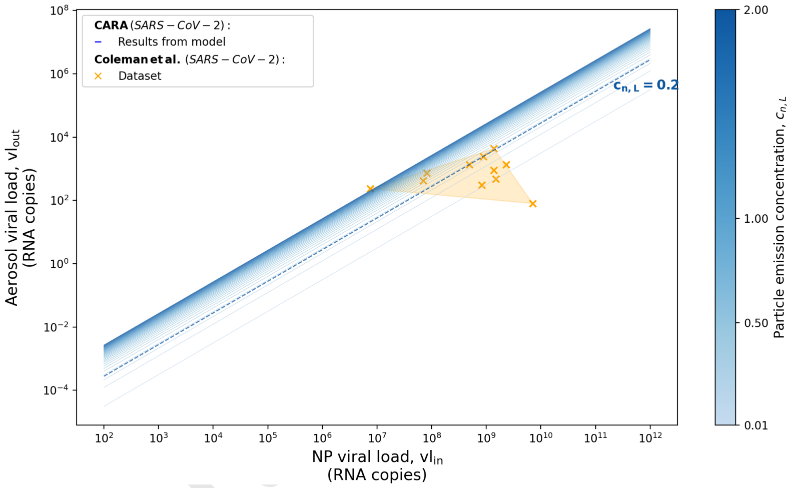


**(b)** Speaking (vocalisation) for 15 minutes

**Figure S. 9:** Exhaled (aerosolized) RNA copies while performing respiratory and vocalisation activities, while seated, without masks, as a function of the viral load from nasopharyngeal (NP) swabs. The blue lines are the result from the model with a degraded color scale ranging the particle emission concentration $c_{n,i}$. The datasets correspond to published clinical trials for influenza [6, 7] and SARS-CoV-2 [8]. a) Results for an infected person breathing for 30 min. b) Results for an infected person under vocal speaking for 15 min. Best fit for the particle emission parameters in the BLO model: $c_{n,B} = 0.06$; $c_{n,L} = 0.2$ cm$^{-3}$. For the SARS-CoV-2 data a Ct to viral load conversion factor was applied following a regression function: $y = 3.095x + 43.69$ [9].

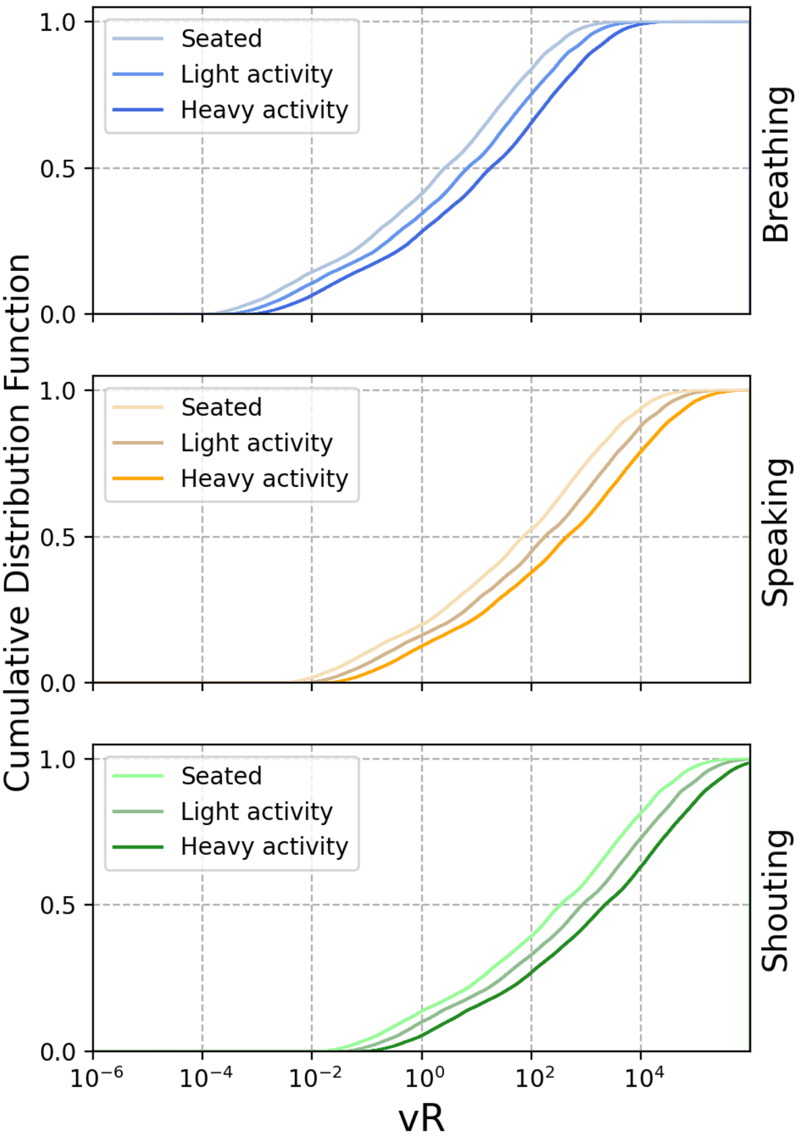


**Figure S. 10:** Estimation of the conditional cumulative probability of vR based on 250 000 MCS for different expiratory activities (Breathing, Speaking and Shouting - from top to bottom) and different physical activities (Seated, Light and Heavy activity). The values are without the effect of face covering ($\eta_{\mathrm{out}} = 0$)

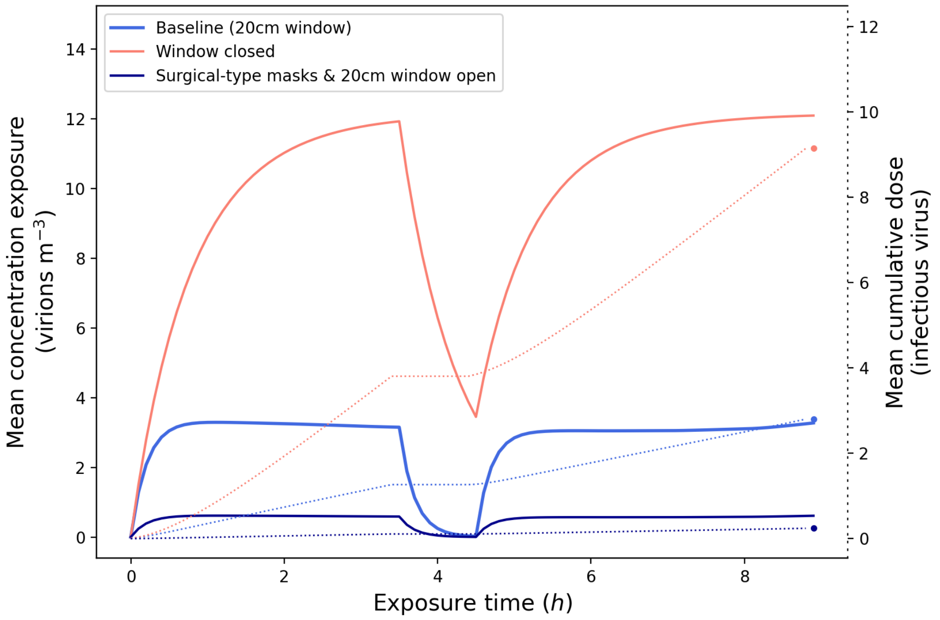


**Figure S. 11:** Results for the viral concentration profile over the exposure time and the cumulative absorbed dose, in the office scenario, for different combination of measures. The solid lines represent the concentration (left y-axis) and the dotted lines represent the cumulative dose (right y-axes). Time = 3.5 corresponds to a 1 hour lunch break. The horizontal section of the dotted lines correspond to the breaks, where the infected and exposed hosts leave the room and are not in contact for the its duration. For visualization purposes, the confidence interval is not represented in the figure. These values can be found in Supplementary Table S4.

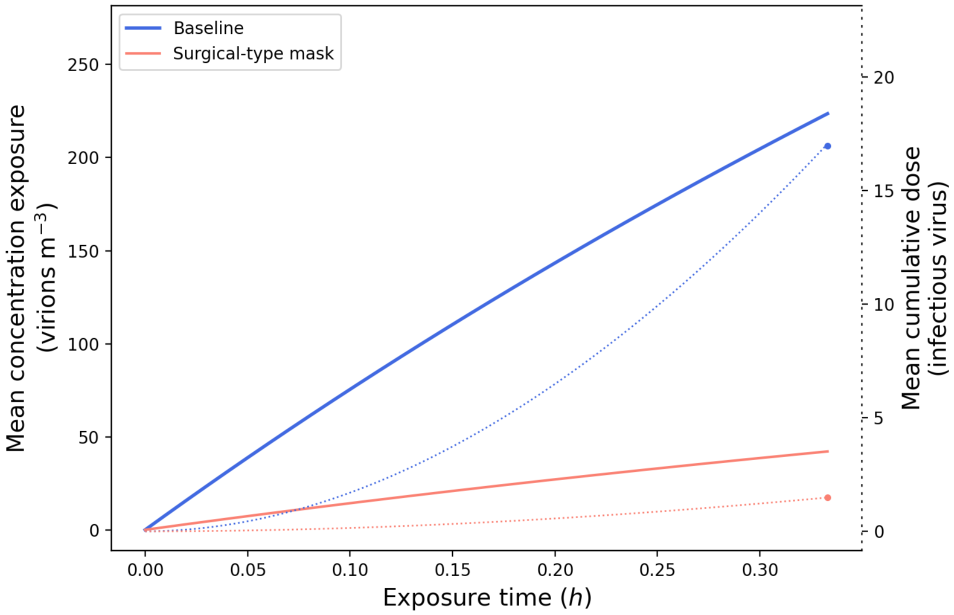


**Figure S. 12:** Results of the viral concentration profile over the exposure time and the cumulative absorbed dose, in the ski cabin scenario, for different combination of measures. The solid lines represent the concentration (left y-axis) and the dotted lines represent the cumulative dose (right y-axes). For visualization purposes, the confidence interval is not represented in the figure. These values can be found in Supplementary Table S4.

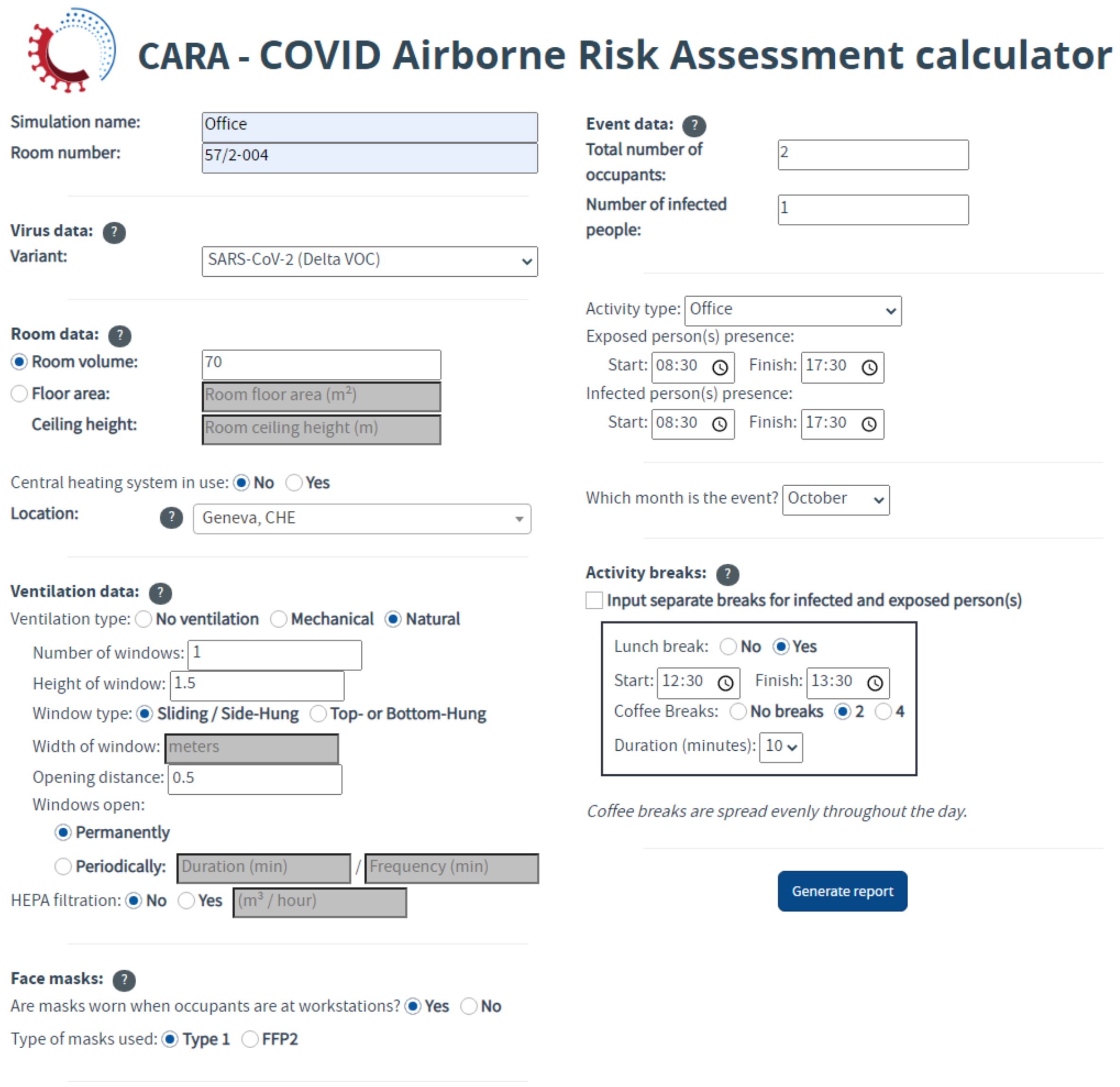


**Figure S. 13:** Screenshot example of the CARA tool input form

Subm

### S.III Tables

**Table S. 1:** Symbol list

| Symbol | Description | Unit | Definition |
|---|---|---|---|
| $D$ | Particle diameter | µm | Section ?? |
| $\mathrm{vR}(D)$ | Emission rate of viruses per unit diameter | virion h$^{-1}$ $\mu$m$^{-1}$ | |
| $E_{c,j}(D)$ | Volumetric particle emission concentration, for each activity *j*, per unit diameter | mL m$^{-3}$ $\mu$m$^{-1}$ | |
| $E_{c,j}^{\mathrm{total}}$ | Total volumetric particle emission concentration, for each activity *j* | mL m$^{-3}$ | |
| $c_{n,i}$ | Total emission concentration | cm$^{-3}$ | |
| $f_{\mathrm{amp}}$ | Vocalisation amplification factor | - | |
| $\mathrm{vl}_{\mathrm{out}}$ | Viral load outside the infected host | RNA copies mL$^{-1}$ | Section ?? |
| $\mathrm{vl}_{\mathrm{in}}$ | Viral load inside the infected host's respiratory track | | |
| $t$ | Duration of the emission, exposure | h | |
| $f_{\mathrm{evap}}$ | Evaporation factor | - | Section ?? |
| $\mu_{D_i}$ | Mean of the natural logarithm of the diameter for each mode $i$ | ln µm | |
| $\sigma_{D_i}$ | Standard deviation of the natural logarithm of the diameter for each mode $i$ | ln µm | |
| $V(D)$ | Volume of the particles for a given diameter $D$ | m$^3$ | |
| $\eta_{\mathrm{out}}$ | Outward mask efficiency | - | Section ?? |
| GM | Geometric mean of the log-normal distribution for the particle diameters | µm | |
| GSD | Geometric standard deviation of the log-normal distribution for the particle diameters | µm | |
| $\mathrm{BR}_{\mathrm{k}}$ | Breathing flow rate for a given physical activity k | m$^3$ h$^{-1}$ | Section ?? |
| $C(t,D)$ | Viral concentration per unit diameter | virion m$^{-3}$ $\mu$m$^{-1}$ | Section ?? |
| $\lambda_{\mathrm{vRR}}$ | Removal rate in the concerned room | h$^{-1}$ | Section ?? |
| $V_r$ | Room volume | m$^3$ | |
| $N_{\mathrm{inf}}$ | Number of infected hosts | - | |
| $\lambda_{\mathrm{vRR}}$ | Viral removal rate | h$^{-1}$ | Section ?? |
| $\lambda_{\mathrm{ACH}}$ | Removal rates related to ventilation | | |
| $\lambda_{\mathrm{dep}}$ | Removal rates related to gravitational settlement | | |
| $\lambda_{\mathrm{bio}}$ | Removal rates related to biological decay | | |
| $\lambda_{\mathrm{HEPA}}$ | Removal rates related to filtration | | |

Submitted for review

| Symbol | Description | Unit | Definition |
|---|---|---|---|
| $Q_{\mathrm{ACH}}$ | Volumetric flow rate of fresh air supplied to the room | $m^3\,h^{-1}$ | Section ?? |
| $C_{\mathrm{d}}$ | Discharge coefficient of the opening | - | |
| $A$ | Area of the opening | $m^2$ | |
| $g$ | Gravitational acceleration | $m\,s^{-2}$ | |
| $h$ | Height of the opening | m | |
| $\Delta T$ | Indoor/outdoor air temperature difference | K | |
| $T_{\mathrm{avg}}$ | Average indoor/outdoor air temperature | K | |
| $L$ | Length of the opening | m | |
| $\phi$ | Window opening angle | $\circ$ | |
| RH | Relative humidity | - | Section ?? |
| $v$ | Settling velocity of a certain particle | $m\,s^{-1}$ | Section ?? |
| $\rho_{\mathrm{p}}$ | Mass density of the airborne particles | $kg\,m^{-3}$ | |
| $\rho_{\mathrm{air}}$ | Mass density of air | $kg\,m^{-3}$ | |
| $D_{\mathrm{evap}}$ | Diameter of the desiccated particle, after evaporation | µm | |
| $\mu_{\mathrm{air}}$ | Dynamic viscosity of air | $kg\,m^{-1}\,s^{-1}$ | |
| $\mathrm{PR}_{20}$ | Particle removal objective | - | Section ?? |
| $Q_{\mathrm{HEPA}}$ | Effective flow rate through the filtering device | $m^3\,h^{-1}$ | |
| $\eta_{\mathrm{f}}$ | Filter efficiency | - | |
| $\mathrm{vD}^{\mathrm{total}}$ | Viral dose | PFU[a] | Section ?? |
| $n$ | Total amount of independent exposures in the same event | - | |
| $f_{\mathrm{inf}}$ | Fraction of infectious virus | - | Section ?? |
| $r_{\mathrm{inf}}$ | Viable-to-RNA virus ratio | - | |
| $\mathrm{HI}_{\mathrm{inf}}$ | Host immunity of the infected population | - | |
| $\eta_{\mathrm{in}}$ | Inward mask efficiency of the PPE | - | Section ?? |
| $f_{\mathrm{dep}}$ | Deposition fraction in the respiratory tract | - | Section ?? |
| $\mathrm{HI}_{\mathrm{exp}}$ | Host immunity of the exposed population | - | Section ?? |
| $P(I)$ | Probability of infection | - | |
| $T_{\mathrm{voc}}$ | Reported increase of transmissibility of a given VOC | - | |
| $\mathrm{ID}_{50}$ | Infectious Dose | - | Section ?? |

[a] The dose can simply be expressed as *infectious viruses* or *viable viruses*

Submitted for review

**Table S. 2:** Statistical data on breathing rate (in $m^3 h^{-1}$) for ages 16-61 (with equal male/female weighting).

| Activity | Mean | Quantiles (%) | | | | | | | |
|---|---|---|---|---|---|---|---|---|---|
| | | 5 | 10 | 25 | 50 | 75 | 90 | 95 | 100 |
| Seated | 0.51 | 0.43 | 0.44 | 0.47 | 0.50 | 0.54 | 0.58 | 0.61 | 0.80 |
| Standing | 0.57 | 0.49 | 0.50 | 0.53 | 0.56 | 0.60 | 0.64 | 0.67 | 0.86 |
| Light Exercise | 1.25 | 1.08 | 1.11 | 1.16 | 1.23 | 1.32 | 1.42 | 1.48 | 1.85 |
| Moderate Exercise | 1.78 | 1.30 | 1.38 | 1.53 | 1.72 | 1.97 | 2.26 | 2.46 | 3.64 |
| Heavy Exercise | 3.30 | 2.28 | 2.45 | 2.78 | 3.17 | 3.71 | 4.30 | 4.75 | 6.73 |

**Table S. 3:** Discharge coefficient parameters for Eq. (**??**) for top- or bottom-hung windows, as a function of the width over height ratio $\frac{w}{h}$ [10]

| | $w/h < 0.5$ | $0.5 \leq w/h < 1$ | $1 \leq w/h < 2$ | $w/h \geq 2$ |
|---|---|---|---|---|
| $C_{\mathrm{d,max}}$ | 0.612 | 0.589 | 0.563 | 0.548 |
| $M$ [deg$^{-1}$] | 0.06 | 0.048 | 0.04 | 0.038 |

**Table S. 4:** Results for the $\mathrm{vD}^{\mathrm{total}}$ distribution (in infectious virus) for the different scenarios and measures.

| **Shared office** | | | |
|---|---|---|---|
| *Scenario* | *Mean* | *5th percentile* | *95th percentile* |
| Baseline | 2.8 | 3.17E-05 | 14.7 |
| W/ masks | 0.2 | 2.56E-06 | 1.3 |
| No ventilation | 9.1 | 1.04E-04 | 46.7 |
| **Classroom** | | | |
| *Scenario* | *Mean* | *5th percentile* | *95th percentile* |
| Baseline | 9.6 | 1.07E-04 | 49.5 |
| Full window open during breaks (winter) | 15.7 | 1.76E-04 | 81.6 |
| Full window open during summer | 5.8 | 6.56E-05 | 30.4 |
| HEPA filter (5 ACH) | 4.5 | 5.12E-05 | 23.3 |
| W/ masks | 0.8 | 8.67E-06 | 4.2 |
| No ventilation | 21.5 | 2.40E-04 | 111.1 |
| **Ski Cabin** | | | |
| *Scenario* | *Mean* | *5th percentile* | *95th percentile* |
| Baseline | 17.0 | 1.83E-04 | 86.8 |
| W/ masks | 1.5 | 1.48E-05 | 7.4 |

Submitted for review

**Table S. 5:** Results for the probability of infection, $P(I)$, and potential number of new (secondary) cases, $N$, for the different scenarios and measures.

| **Shared office** | | | | | | |
|---|---|---|---|---|---|---|
| *Scenario* | *P(I) (Mean)* | *P(I) (5th per.)* | *P(I) (95th per.)* | *Occupants (exposed)* | *N (mean)* | *N (95th per.)* |
| Baseline | 0.06 | 8E-07 | 0.37 | 3 | 0.18 | 1.10 |
| W/ masks | 0.01 | 7E-08 | 0.04 | | 0.02 | 0.11 |
| No ventilation | 0.13 | 3E-06 | 0.78 | | 0.38 | 2.33 |
| **Classroom** | | | | | | |
| *Scenario* | *P(I) (Mean)* | *P(I) (5th per.)* | *P(I) (95th per.)* | *Occupants (exposed)* | *N (mean)* | *N (95th per.)* |
| Baseline | 0.13 | 3E-06 | 0.78 | 19 | 2.4 | 14.9 |
| Full window open during breaks (winter) | 0.16 | 5E-06 | 0.92 | | 3.1 | 17.5 |
| Full window open during summer | 0.10 | 2E-06 | 0.66 | | 2.0 | 12.6 |
| HEPA filter (5 ACH) | 0.08 | 1E-06 | 0.52 | | 1.6 | 9.9 |
| W/ masks | 0.02 | 2E-07 | 0.12 | | 0.4 | 2.3 |
| No ventilation | 0.19 | 7E-06 | 0.97 | | 3.6 | 18.4 |
| **Ski Cabin** | | | | | | |
| *Scenario* | *P(I) (Mean)* | *P(I) (5th per.)* | *P(I) (95th per.)* | *Occupants (exposed)* | *N (mean)* | *N (95th per.)* |
| Baseline | 0.17 | 5E-06 | 0.93 | 3 | 0.5 | 2.8 |
| W/ masks | 0.04 | 4E-07 | 0.21 | | 0.1 | 0.6 |
| **Outbreaks (benchmark)** | | | | | | |
| *Scenario* | *P(I) (Mean)* | *P(I) (5th per.)* | *P(I) (95th per.)* | *Occupants (exposed)* | *N (mean)* | *N (95th per.)* |
| SV Chorale[a] | 0.7 | 0.4 | 0.8 | 60 | 42.5 | 49.2 |
| Bus ride[a] | 0.1 | 0.04 | 0.3 | 67 | 8.0 | 20.7 |

[a] Assuming $\mathrm{vl}_{\mathrm{in}}$ is $10^9$ and $5 \cdot 10^8$ copies per mL for the SV Chorale and Bus ride index hosts, respectively.

Submitted for review

**Table S. 6:** Data gathered from Mikszewski et al [11] (first 3 columns) and converted to reflect the units of vR

| **Virus** | **Setting** | **quanta / h** | **virion / h** (lower bound) | **virion / h** (upper bound) |
|---|---|---|---|---|
| SARS-CoV | Hospital | 29 | 418 | 4176 |
| SARS-CoV-2 | Apartment | 15 | 216 | 2160 |
| SARS-CoV-2 | Ship | 15 | 216 | 2160 |
| SARS-CoV-2 | Bus | 36 | 518 | 5184 |
| SARS-CoV-2 | Bus ride (benchmark scenario) | 45 | 648 | 6480 |
| SARS-CoV-2 | Restaurant | 61 | 878 | 8784 |
| SARS-CoV-2 | Bus | 62 | 893 | 8928 |
| SARS-CoV-2 | School | 116 | 1670 | 16704 |
| SARS-CoV-2 | Courtroom | 130 | 1872 | 18720 |
| SARS-CoV-2 | Bus | 133 | 1915 | 19152 |
| SARS-CoV-2 | School | 139 | 2002 | 20016 |
| SARS-CoV-2 | Meeting room | 139 | 2002 | 20016 |
| SARS-CoV-2 | Gym | 152 | 2189 | 21888 |
| SARS-CoV-2 | Abattoir | 232 | 3341 | 33408 |
| SARS-CoV-2 | Call Center | 683 | 9835 | 98352 |
| SARS-CoV-2 | S V Chorale (benchmark scenario) | 970 | 13968 | 139680 |
| SARS-CoV-2 | Choir | 4213 | 60667 | 606672 |
| Measles | Classroom | 18 | 259 | 2592 |
| Measles | Classroom | 600 | 8640 | 86400 |
| Measles | Classroom | 2765 | 39816 | 398160 |
| Measles | Office | 8640 | 124416 | 1244160 |
| Influenza | Quarantine rooms | 0.11 | 2 | 16 |
| Influenza | Clinical tria (ferrets) | 7.95 | 114 | 1145 |
| Influenza | Airplane | 79 | 1138 | 11376 |
| Rhinovirus | Clinical trial (humans, leisure) | 3.1 | 45 | 446 |

Submitted for review


Submitted for review
